# Shared additive- and variance-associated genetic architecture of cardiac structure and function

**DOI:** 10.64898/2026.11.04.26367193

**Authors:** James Tsaggaris, Nour Mimouni, Renae L. Judy, Diane Xue, Michael P. Morley, Xiao Wang, Justin L. Cotney, Daniel P. Kelly, Sharlene M. Day, Thomas P. Cappola, Penn Medicine BioBank, Scott M. Damrauer, Michael G. Levin

## Abstract

Cardiac remodeling underlies cardiomyopathy and heart failure, yet how common genetic effects are organized across the four chambers, and whether they influence phenotypic variability beyond additive genetic effects, remains unclear. We analyzed 22 cardiac magnetic resonance phenotypes in up to 77,271 UK Biobank participants. Additive mapping identified 748 locus–trait associations across 239 regions. Multivariate fine-mapping revealed that 96% of signals showed effects across multiple cardiac chambers, while resolving independent signals with distinct anatomical profiles, including predominantly atrial and ventricular signals at RBM20. Fine-mapped signal sharing formed three robust communities corresponding broadly to atrial function, chamber remodeling and ventricular systolic function. Variance mapping identified 19 associations, 13 without a nearby additive signal. Individual variance loci and polygenic effects generalized to clinical echocardiographic phenotypes and outcomes in the Penn Medicine Biobank. These findings reveal cardiac common-variant architecture as broadly shared yet locally heterogeneous and identify phenotypic variability as a partly distinct genetic dimension.

## Introduction

Cardiomyopathies and heart failure are characterized by remodeling of cardiac structure and function^1^. Recent studies leveraging cardiac magnetic resonance (CMR) in large cohorts have demonstrated that cardiac structure and function are heritable and have identified common variants influencing CMR-derived phenotypes, including loci near genes implicated in cardiac development and cardiomyopathy^2–5^. Polygenic variation associated with CMR traits has also been associated with incident cardiomyopathy and with heart failure, supporting a link between common genetic influences on quantitative cardiac phenotypes and clinical cardiovascular disease^2–5^.

Prior CMR GWAS have identified genetic determinants of cardiac phenotypes and demonstrated substantial genetic correlation and pleiotropy across cardiac traits^2–4^. However, overlap at the locus level does not establish whether the same underlying genetic signal influences multiple phenotypes or whether distinct effects coexist within the same region^6^. The extent to which distinct signals within cardiac loci contribute to single or multiple phenotypes, the genes underlying these associations, and their relationships with cardiomyopathy and heart failure remain incompletely understood.

These studies have primarily characterized genetic effects on mean phenotype levels, whereas genetic influences on phenotypic variability remain largely unexplored^2–4^, limiting our understanding of context-dependent genetic effects. Although cardiac phenotypes can show substantial heterogeneity among individuals carrying pathogenic cardiomyopathy variants^7^, exhaustively testing all possible gene–gene or gene–environment interactions requires the interacting factors to be specified and is impractical at the genome scale. Variance quantitative trait locus (vQTL) mapping instead identifies variants associated with differences in phenotypic variance and can therefore prioritize loci at which genetic effects may depend on genetic or environmental context^8^. To our knowledge, vQTL mapping has not previously been applied to CMR-derived traits.

We performed genome-wide additive and variance association analyses of 22 CMR-derived phenotypes spanning all four cardiac chambers in up to 77,271 UK Biobank participants. We sought to determine whether common genetic effects on cardiac structure and function are broadly shared across phenotypes and chambers or exhibit anatomically distinct patterns; whether loci underlying quantitative CMR traits converge on genes and genetic signals implicated in cardiomyopathy and heart failure; and whether common variants also influence phenotypic variability beyond their additive effects on trait levels. To validate our findings, we further evaluated whether additive and variance-associated polygenic effects generalized to clinically acquired cardiac phenotypes in Penn Medicine Biobank.

## Results

### Genetic associations are extensively shared across cardiac phenotypes

In total, 77,271 UK Biobank participants were included. For these individuals, chamber measurements were derived by deep-learning segmentation^9^, yielding measurements across the cardiac chambers. These comprised left atrial (LA) ejection fraction (LAEF), minimum volume (LAMIV), maximum volume (LAMV) and stroke volume (LASV); left ventricular (LV) ejection fraction (LVEF), end-diastolic volume (LVEDV), end-systolic volume (LVESV), stroke volume (LVSV), cardiac output (LVCO), myocardial mass (LVM), global mean myocardial wall thickness (LVWTG), global longitudinal strain (LVLSG), global circumferential strain (LVCSG) and global radial strain (LVRSG); right atrial (RA) ejection fraction (RAEF), minimum volume (RAMIV), maximum volume (RAMV) and stroke volume (RASV); and right ventricular (RV) end-diastolic volume (RVEDV), ejection fraction (RVEF), end-systolic volume (RVESV) and stroke volume (RVSV).

We performed additive GWAS for each phenotype for 10.7 million variants with minor allele frequency (MAF) ≥ 0.005 and imputation score (INFO) > 0.8. Single nucleotide polymorphism (SNP)-based heritability estimates (*h*^2^) ranged from 6.7% (LAEF) to 24.0% (RVESV), with a median of 13.5% (Supplementary Table 1). The highest estimates were observed for ventricular phenotypes, including RVESV (24.0%), RVEDV (23.6%), LVEDV (22.2%), LVM (21.7%), LVWTG (19.4%), and LVESV (19.3%). Across 22 CMR phenotypes, we identified 748 genome-wide significant locus–trait associations, corresponding to 239 unique genomic regions (Fig. 1a–c; Supplementary Table 2).

**Fig. 1:**
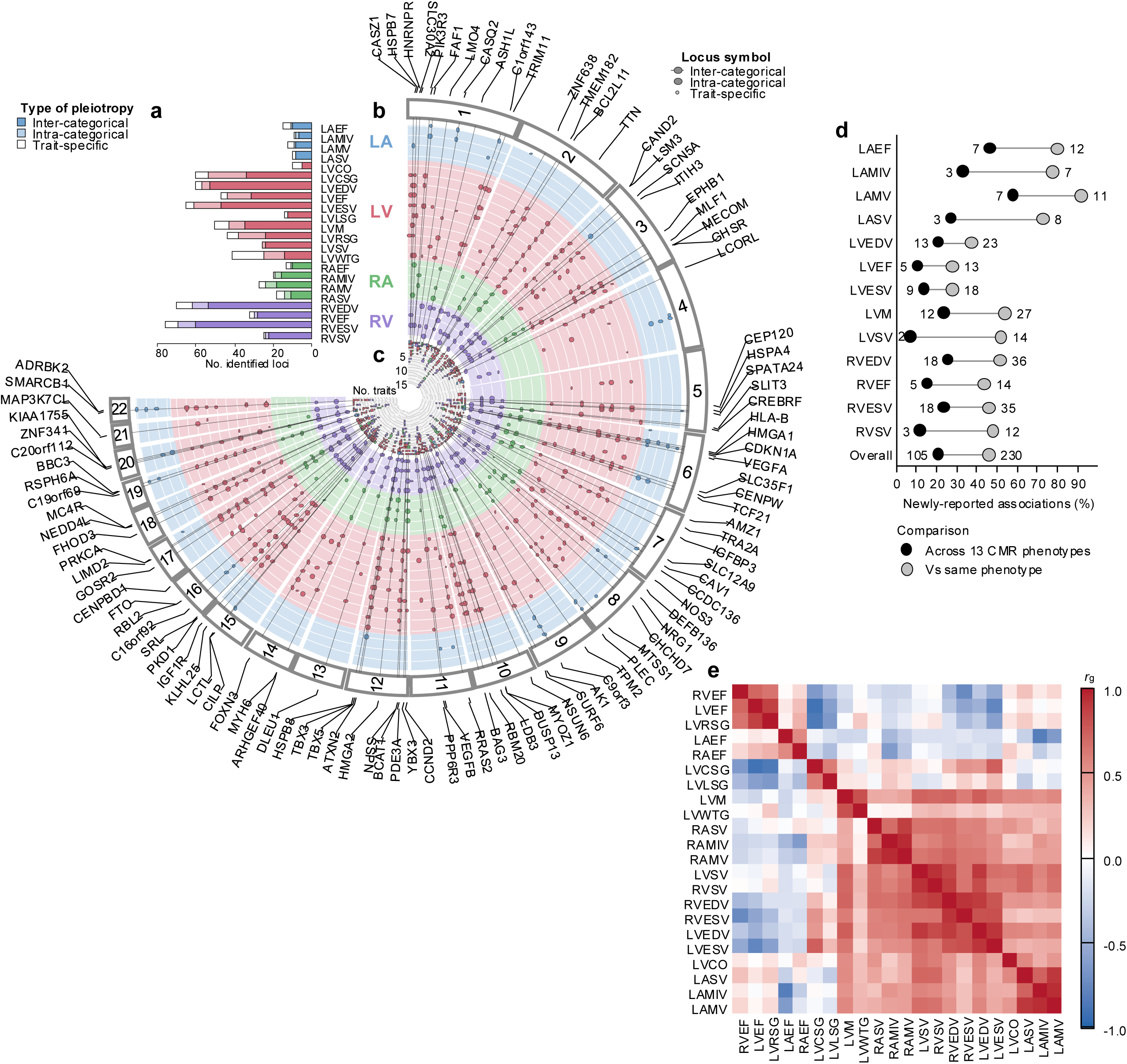
Additive genetic associations are extensively shared across cardiac phenotypes. **a**, Number of genome-wide significant loci identified for each of 22 cardiac magnetic resonance (CMR) phenotypes, classified as inter-categorical, intra-categorical or trait-specific according to locus sharing across cardiac chambers. Colors denote left atrial (LA), left ventricular (LV), right atrial (RA) and right ventricular (RV) phenotypes. **b**, Circular representation of genome-wide significant additive associations across the 22 CMR phenotypes. Points denote locus–trait associations. Nearest genes are labeled for inter-categorical loci. **c**, Concentric scale indicating the number of CMR phenotypes associated with each genomic region. **d**, Percentage of locus–trait associations newly reported relative to previous CMR genome-wide association studies, assessed either across all 13 comparable CMR phenotypes (black circles) or relative to the same phenotype (gray circles). Numbers indicate the corresponding counts of newly reported associations. **e**, Pairwise genetic correlations (*rg*) among the 22 CMR phenotypes estimated using LD Score Regression. Color indicates the magnitude and direction of *rg*. Additive association analyses included up to 77,271 UK Biobank participants.

To determine which genomic regions had not been previously identified, we mapped our traits to findings from prior CMR GWAS. We identified 13 phenotypes corresponding to recent large CMR GWAS^2–4^. Among those 13 traits, we identified 501 genome-wide significant locus– trait associations. A locus was classified as novel relative to the included studies when no previously reported genome-wide significant variant was located within 2 Mb of either locus boundary. Among these associations, 230 (45.9%) were novel relative to the same phenotype, and 105 (21.0%) were novel across all of the 13 comparable phenotypes (Fig. 1d; Supplementary Table 2). Notably, 125 (25.0%) loci had previously been associated with a different CMR phenotype, highlighting extensive locus-level overlap across cardiac traits. However, such overlap does not establish whether the same underlying genetic signal influences multiple phenotypes or whether distinct effects coexist within the same region.

### Multivariate analyses resolve genetic effects across cardiac phenotypes

To evaluate the degree of shared common genetic variation influencing CMR traits, we examined the genetic signals at both the global- and locus-levels. Pairwise LD score regression (LDSC)^10^ revealed extensive genetic correlation between phenotypes (Fig. 1e; Supplementary Table 3). Among 231 pairs (median |*r_g_*| = 0.341), 181 (78.4%) were significant (FDR < 0.05; Supplementary Table 3), and the strongest genetic correlations reflected biologically related phenotypes, including traits within the same chamber (RAMIV–RAMV), homologous traits across chambers (LVSV–RVSV), and related structural measures (LVM–LVWTG). These genome-wide correlations establish widespread genetic sharing but do not determine whether associations within individual loci arise from the same or distinct underlying genetic effects.

To resolve the individual genetic signals influencing the CMR traits, we then performed multivariate fine-mapping using mvSuSiE^6^ across the additive regions for 18 cardiac traits. Because the four stroke-volume traits (LASV, LVSV, RASV and RVSV) are derived from the corresponding volumetric measures, we excluded them to avoid including highly correlated traits. We identified 669 credible sets (CS) (Supplementary Table 4), all of which demonstrated effects across multiple phenotypes (Supplementary Fig. 1), with 644 showing effects across multiple cardiac chambers. Nearly half of the fine-mapped signals showed effects across all four chambers (330, 49.3%), while 141 (21.1%) showed effects restricted to the left and right ventricles (Supplementary Fig. 2). Most fine-mapped signals therefore showed effects across multiple chambers, although the breadth of sharing at each locus varied substantially (Supplementary Table 5).

To examine higher-order organization of this sharing, we represented mvSuSiE credible sets and the phenotypes they supported as a bipartite network and applied community detection directly to the observed network. Three phenotype communities emerged (Barber bipartite modularity^11^ *Q* = 0.181; Supplementary Table 6). One community comprised the atrial ejection-fraction traits LAEF and RAEF; a second community grouped atrial volume measures with LVEDV, RVEDV, LVCO, LVM and LVWTG, corresponding broadly to chamber size and remodeling; and a third community comprised ventricular ejection fraction, end-systolic volumes and LV strain measures, corresponding broadly to ventricular systolic function (Supplementary Table 6). The observed modularity exceeded that of 10,000 randomized networks preserving both credible-set and phenotype degree distributions (empirical *P* = 1.0 × 10^−4^), and the same phenotype partition was recovered in all 10,000 random reorderings of the observed network (Supplementary Table 7).

Complementing this data-driven organization, we tested whether credible-set sharing followed prespecified anatomical groupings (Supplementary Table 8). Phenotypes within the same atrial or ventricular compartment shared a greater fraction of credible sets than atrial–ventricular phenotype pairs (mean Jaccard similarity, 47.2% versus 35.4%; permutation *P* = 0.0028), whereas no corresponding organization was observed by left–right laterality or individual cardiac chamber (*P* = 0.69 and *P* = 0.13, respectively; Supplementary Table 9 and Supplementary Fig. 3). Sharing was particularly extensive between corresponding ventricular measures, including LVEDV– RVEDV (83.6%), LVESV–RVESV (81.5%) and LVEF–RVEF (67.0%) (Supplementary Fig. 4).

At individual loci, shared and anatomically restricted effects could coexist. A single-variant signal at the *IGF1R* locus showed effects across all 18 analyzed phenotypes, illustrating the broad phenotypic effects observed at some loci (Fig. 2a). At the *RBM20* locus, mvSuSiE resolved six distinct signals with differing patterns of phenotypic sharing (Fig. 2b). A single-variant signal led by 10:112544125:C:T (rs189569984; PIP = 1.00) showed effects across 10 left- and right-ventricular phenotypes; the same missense variant was previously associated with LVESV, RVESV and LVEF in a UK Biobank exome-imaging analysis^12^. An independent single-variant signal led by 10:112576695:T:C (rs10749053; PIP = 0.977) showed a predominantly atrial pattern; rs10749053 was previously identified as an atrial-fibrillation risk variant^13^. The region encompassing *TTN* and *CCDC141* showed the greatest signal complexity, with eight credible sets, including four single-variant signals. One single-variant signal showed effects across 16 phenotypes spanning all four cardiac chambers, whereas another showed effects only in 10 left-and right-ventricular phenotypes (Fig. 2c). Two of the signals were led by missense variants in *CCDC141*: p.Arg935Trp (2:179721046:G:A; PIP = 0.512; two-variant credible set) and p.Ala141Val (2:179839888:G:A; PIP = 1.00), both showing effects across all four chambers. These same variants were previously identified as independent genome-wide significant risk variants for accessory atrioventricular pathways and were also associated with paroxysmal supraventricular tachycardia^14^. Together, these examples illustrate that extensive cardiac pleiotropy can coexist with marked signal-level specificity within the same genomic region.

**Fig. 2:**
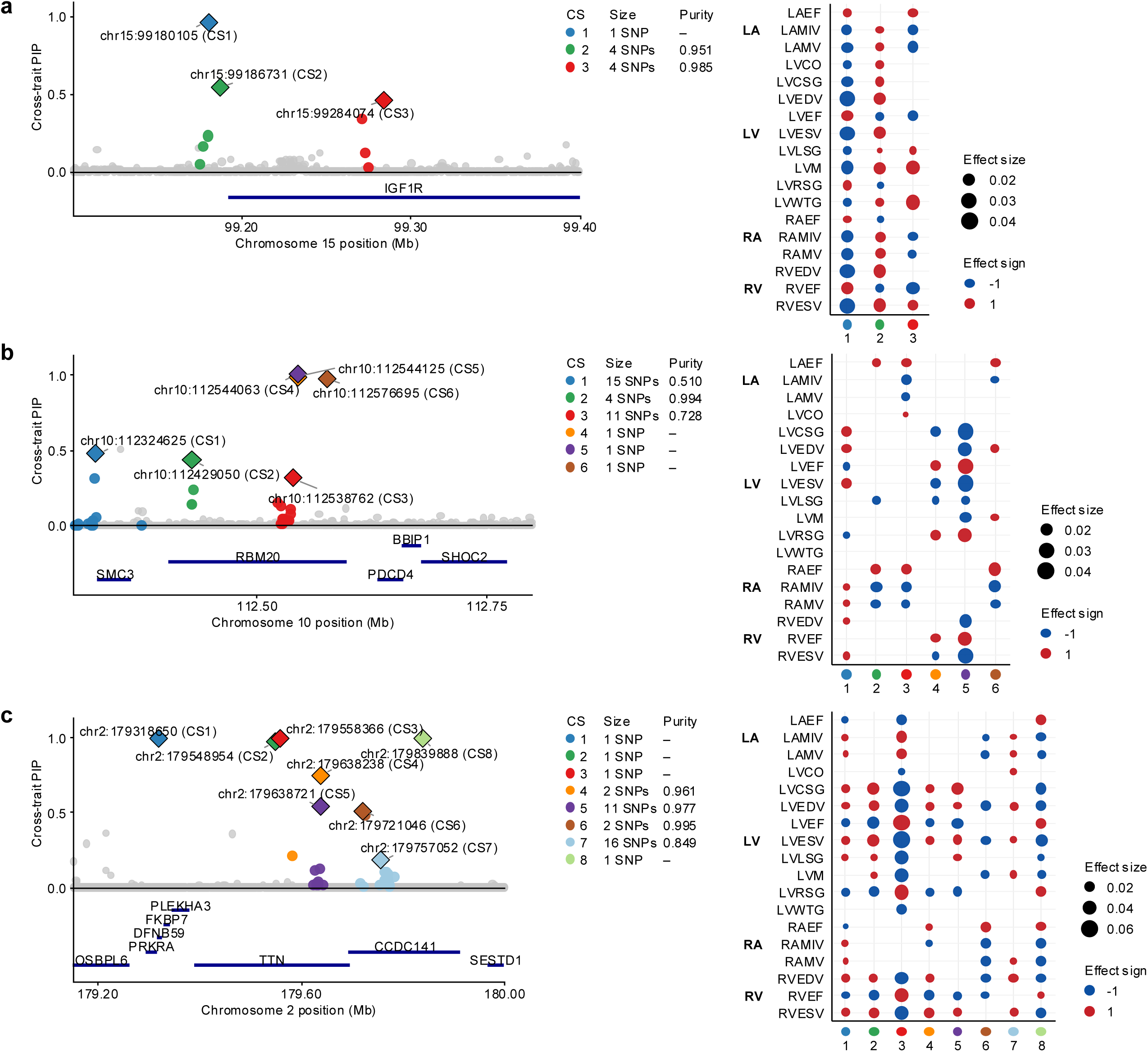
Multivariate fine-mapping resolves shared and anatomically differentiated signals within cardiac loci. **a–c**, Multivariate fine-mapping with mvSuSiE at the *IGF1R* (**a**) and *RBM20* (**b**) loci and the *TTN–CCDC141* region (**c**). Left, variant-level cross-trait posterior inclusion probabilities (PIPs); colors identify distinct credible sets (CSs), and diamonds indicate the variant with the highest PIP within each CS. Credible-set size and purity are indicated; purity is not applicable to single-variant credible sets. Right, phenotypic support for each credible set across the 18 CMR phenotypes included in multivariate fine-mapping. Circle size indicates the magnitude of the conditional effect and color indicates effect direction (red, positive; blue, negative). Dots are shown for phenotypes supported at a local false sign rate ≤0.01.

### Cardiac genetic signals converge on developmental and cardiomyocyte biology

We next investigated the biological context of these associations through gene prioritization and enrichment analyses. Single-trait fine-mapping of the 748 additive locus–trait associations yielded 650 credible sets, with at least one credible set identified for 594 associations. Of these, 36 were single-variant credible sets and 135 contained ≤ 5 variants. We integrated fine-mapping and gene-level evidence using FLAMES^15^ to prioritize candidate genes for each credible set. High-confidence genes (cumulative precision > 75%) were prioritized at 304 credible sets, corresponding to 149 unique genes (Supplementary Table 10). Recurrently prioritized genes included *TTN* (11 CS across 6 traits), *TBX3* (6 CS across 6 traits), *GJA1* and *RBM20* (4 CS across 4 traits each), and *PITX2* (5 CS across 3 traits).

Gene Ontology enrichment^16,17^ of pooled CMR-prioritized genes highlighted biological processes related to cardiac development and morphogenesis, including heart morphogenesis, ventricular septum morphogenesis, and ventricular cardiac muscle tissue morphogenesis (Fig. 3a; Supplementary Table 11).

**Fig. 3:**
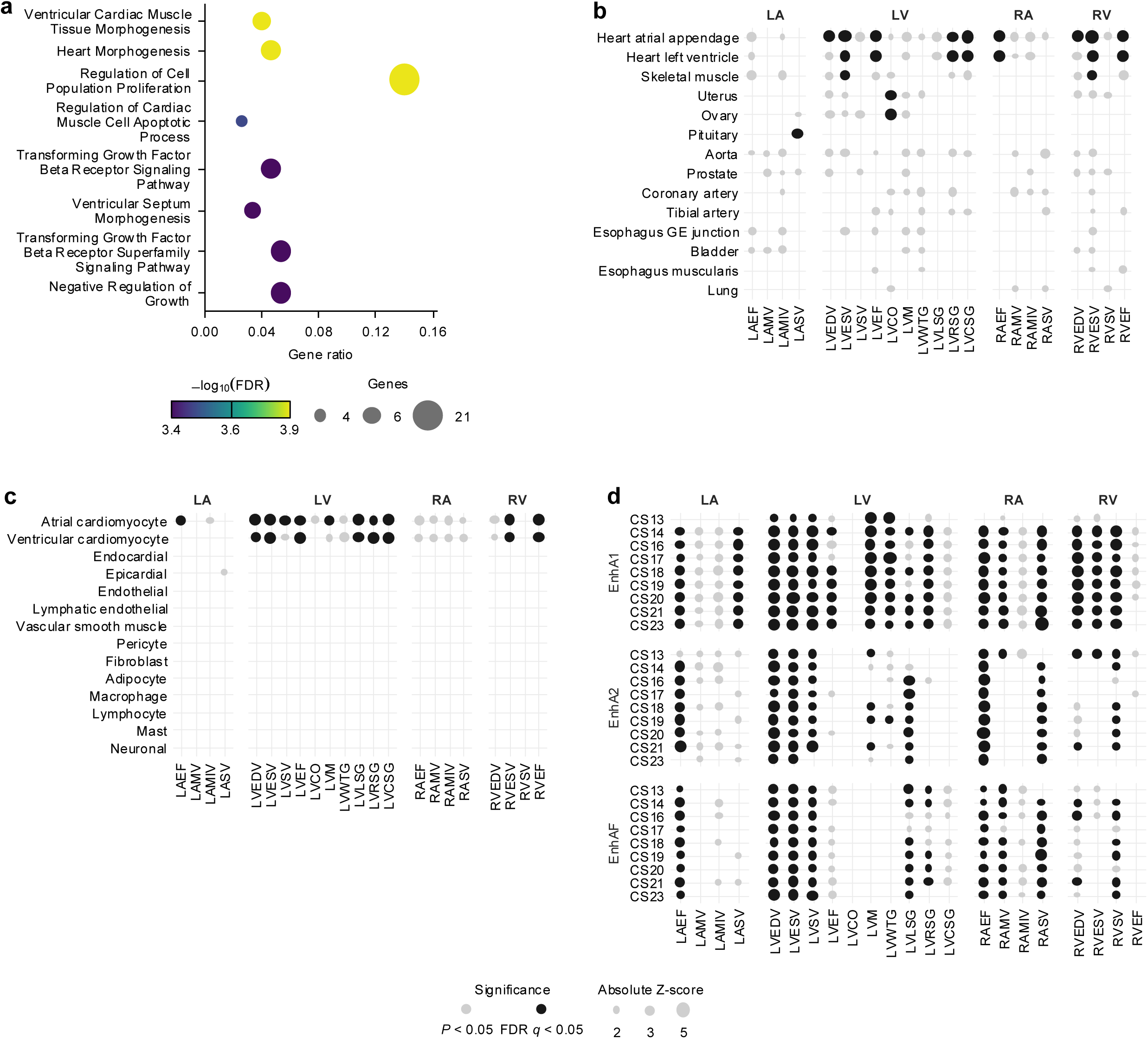
Cardiac genetic associations converge on developmental programs and cardiomyocyte biology. **a**, Gene Ontology Biological Process enrichment among high-confidence genes prioritized from additive CMR associations. Gene ratio indicates the proportion of genes in the prioritized set represented in each term; circle size indicates the number of genes and color indicates −log_10_(FDR-adjusted P value). **b**, Stratified LD Score Regression enrichment across GTEx tissue annotations for the 22 CMR phenotypes. **c**, Enrichment across cardiac cell-type annotations derived from the HeartMap single-nucleus atlas. **d**, Enrichment in active developmental cardiac enhancer annotations across Carnegie stages (CS) and the EnhA1, EnhA2 and EnhAF enhancer states. In **b–d**, phenotypes are grouped by cardiac chamber; black circles indicate FDR q < 0.05, gray circles indicate nominal P < 0.05 and circle size represents the absolute Z score. FDR correction was performed across all annotations within each phenotype and analysis.

Separately, stratified LD Score Regression^18^ using GTEx annotations^19^ showed false discovery rate (FDR)-significant enrichment in atrial appendage and left ventricular tissue for 9 and 7 traits, respectively, accounting for 16 of 21 significant trait–tissue enrichments. Additional signals were found in several non-cardiac tissues, including skeletal muscle and uterus (Fig. 3b; Supplementary Table 12). HeartMap annotations derived from an adult human cardiac single-nucleus RNA-sequencing atlas^20^ further localized this signal to cardiomyocytes, with atrial and ventricular cardiomyocyte enrichment across 11 and 8 traits, respectively, and no other cell type significant after FDR correction (Fig. 3c; Supplementary Table 13).

We also examined regulatory elements active during cardiac development using stratified LD Score Regression of additive GWAS summary statistics and stage-specific enhancer annotations from embryonic human hearts in the Heart Hub^21^. We observed broad heritability enrichment across active enhancer states and multiple stages of cardiac organogenesis. In total, 16 of 22 traits showed FDR-significant enrichment, with signals spanning all nine assayed Carnegie stages and all three active enhancer states. Enrichment was most prominent in the EnhA1 active-enhancer state and was particularly widespread for LVEDV and LVESV, each showing FDR-significant enrichment in 51 of 54 developmental enhancer annotations (Fig. 3d; Supplementary Table 14). Together, these analyses linked common genetic variation in adult cardiac structure and function to both mature cardiomyocyte biology and regulatory programs active during cardiac development.

### Cardiac genetic signals converge on cardiomyopathy and heart failure

We next examined whether quantitative CMR traits shared genetic determinants with pathological cardiac remodeling. CMR-prioritized genes were enriched for established dilated cardiomyopathy (DCM) genes^7^ (3 overlaps: *RBM20*, *SCN5A*, and *TTN*; 7.49-fold enrichment, *P* = 0.0061) and for the combined cardiomyopathy gene set of established hypertrophic cardiomyopathy (HCM)^7^, DCM^7^, and arrhythmogenic cardiomyopathy^22^ genes (5.88-fold enrichment, *P* = 0.0039; Supplementary Table 15). Enrichment was not significant for HCM alone, and no overlap was observed with the curated arrhythmogenic cardiomyopathy gene set.

At the signal level, we tested colocalization between CMR associations and genetic signals for HCM^23^, DCM^24^, and heart failure^25^ established in prior GWAS. Across 239 merged regions, 9 (3.8%) contained at least one high-probability (HyPrColoc^26^ posterior probability > 0.7) CMR– disease colocalization cluster, including 3 regions with HCM, 5 with DCM and 4 with heart failure (Supplementary Table 16). One region, at the BAG3 locus, contained a cluster involving all three diseases; variation at BAG3 has previously been reported to modify risk in opposite directions for DCM and HCM^27^. These findings indicate that a subset of loci influencing quantitative cardiac structure and function also harbor shared genetic signals across the cardiomyopathy disease spectrum.

Several loci showed convergence across multivariate fine-mapping, gene prioritization and disease colocalization. For example, at the *TMEM182* locus, mvSuSiE resolved 2:103426177:C:T (rs2540277) as a single-variant credible set (PIP > 0.999), showing effects across nine left-and right-ventricular phenotypes (Fig. 4a). The variant lies within an intron of the MANE Select *TMEM182* transcript and was also identified by HyPrColoc for a cluster comprising LVCSG, LVEF, LVESV and LVRSG together with HCM and DCM (posterior probability = 0.915). *TMEM182* was also prioritized for both LVCSG and LVRSG. Notably, rs2540277 was itself the lead variant at this locus in the prior HCM GWAS (OR = 1.32, *P* = 2.31 × 10^−8^)^23^ and prior DCM genetic analyses implicated *TMEM182* through LV eQTL colocalization, with lower genetically predicted LV expression associated with increased DCM risk^28^.

**Fig. 4:**
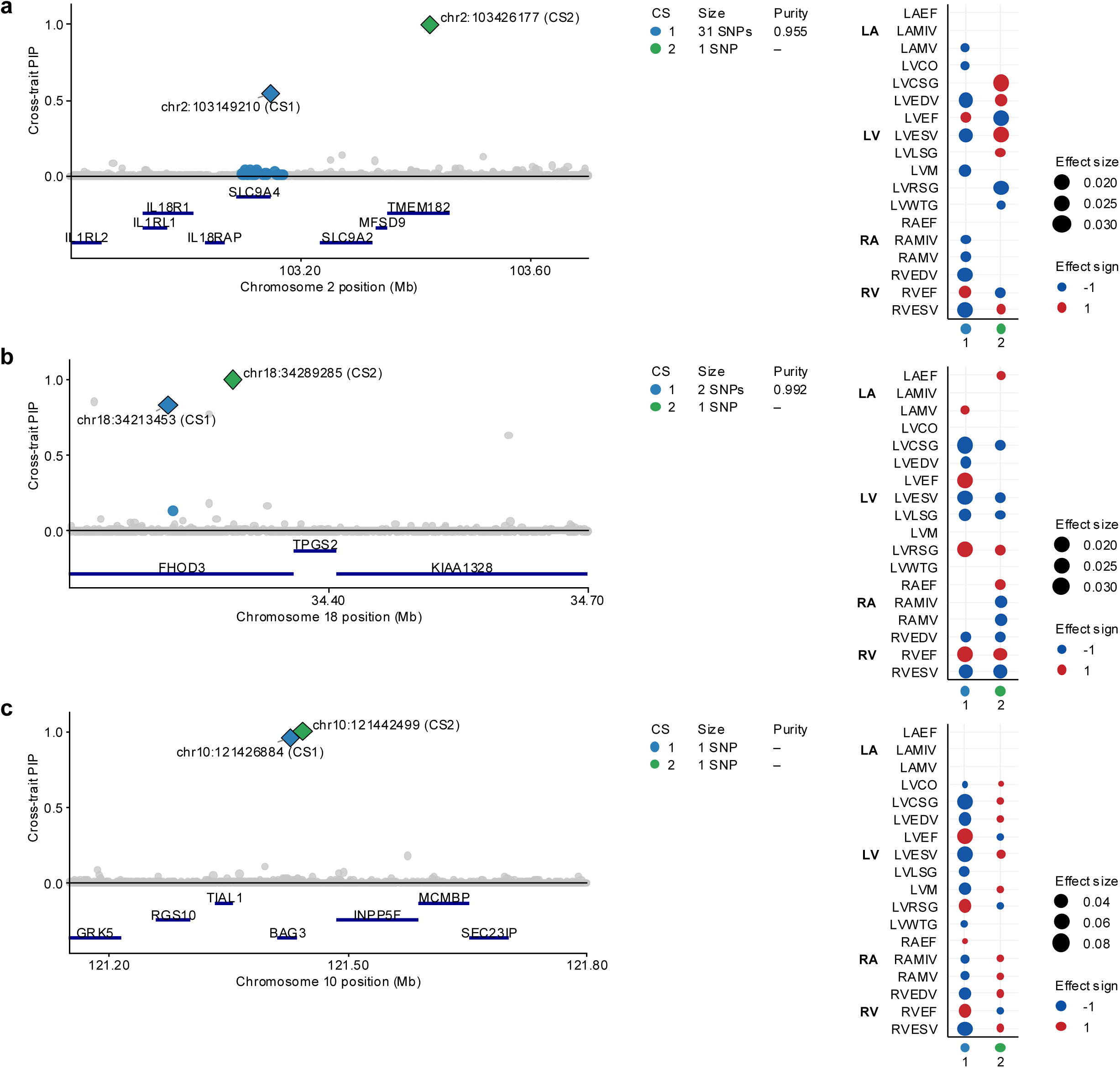
Multivariate fine-mapping resolves cardiac signals at loci shared with cardiomyopathy and heart failure. **a–c**, Multivariate fine-mapping at the *TMEM182* (**a**), *FHOD3* (**b**) and *BAG3*/*INPP5F* (**c**) loci, which contained CMR signals colocalizing with cardiomyopathy or heart failure. Left, variant-level cross-trait posterior inclusion probabilities (PIPs); colors distinguish credible sets (CSs), diamonds indicate the highest-PIP variant within each CS and gene positions are shown below. Credible-set size and purity are indicated, with purity not applicable to single-variant credible sets. Right, phenotypic support for each credible set across the 18 CMR phenotypes. Circle size indicates the magnitude of the conditional effect and color indicates effect direction (red, positive; blue, negative). Dots are shown for phenotypes supported at a local false sign rate ≤0.01.

At the *FHOD3* locus, mvSuSiE resolved two distinct signals. The top variant of a two-variant credible set (18:34213453:A:G; PIP = 0.827) was intronic in *FHOD3* and was also the candidate variant for a DCM colocalization cluster spanning multiple left- and right-ventricular phenotypes (posterior probability = 0.857). This variant was in strong linkage disequilibrium with rs2644262 (Pan-UKB EUR r^2^ = 0.89), the lead *FHOD3*/*TPGS2* variant in the prior HCM GWAS^23^. An independent single-variant signal (18:34289285:G:T; PIP > 0.999) was a missense variant in the MANE Select *FHOD3* transcript (p.Val822Phe; rs61735998) and showed effects across 11 phenotypes spanning all four chambers (Fig. 4b). The same missense variant was previously assigned 100% posterior probability of driving a resting-heart-rate association signal in annotation-informed fine-mapping^29^. *FHOD3* was independently prioritized across three single-trait credible sets from two traits.

At a chromosome 10 locus, 10:121426884:C:T (rs17617337) was resolved as a single-variant credible set (PIP = 0.958) and was the candidate variant for a colocalization cluster spanning 13 CMR phenotypes together with HCM, DCM and heart failure (posterior probability = 0.776) (Fig. 4c). rs17617337 was previously identified as a heart-failure sentinel variant and is in near-perfect linkage disequilibrium with the *BAG3* missense variant p.Cys151Arg (rs2234962; r^2^ = 0.99)^30^, which was the lead BAG3 variant in the prior HCM GWAS^23^. The variant lies within an intron of the MANE Select *BAG3* transcript, whereas integrated gene prioritization independently implicated *INPP5F* across three single-trait credible sets for LVEF and LVCSG. These loci illustrate how genetic effects on quantitative cardiac structure and function can converge with disease-associated signals while retaining substantial allelic and phenotypic complexity.

### Variance QTLs identify genetic effects on cardiac phenotypic variability

All analyses thus far characterized additive genetic effects on CMR phenotypes. We next asked whether common variants also influence interindividual phenotypic variability. Genome-wide variance quantitative trait locus mapping analyses using QUAIL^8^ identified 19 significant locus– trait associations across 14 CMR phenotypes (Fig. 5a–c; Supplementary Table 17). Of these, 13 (68.4%) had no genome-wide significant additive signal within 500 kb, indicating that most variance-associated loci were not captured by nearby additive-trait associations.

**Fig. 5:**
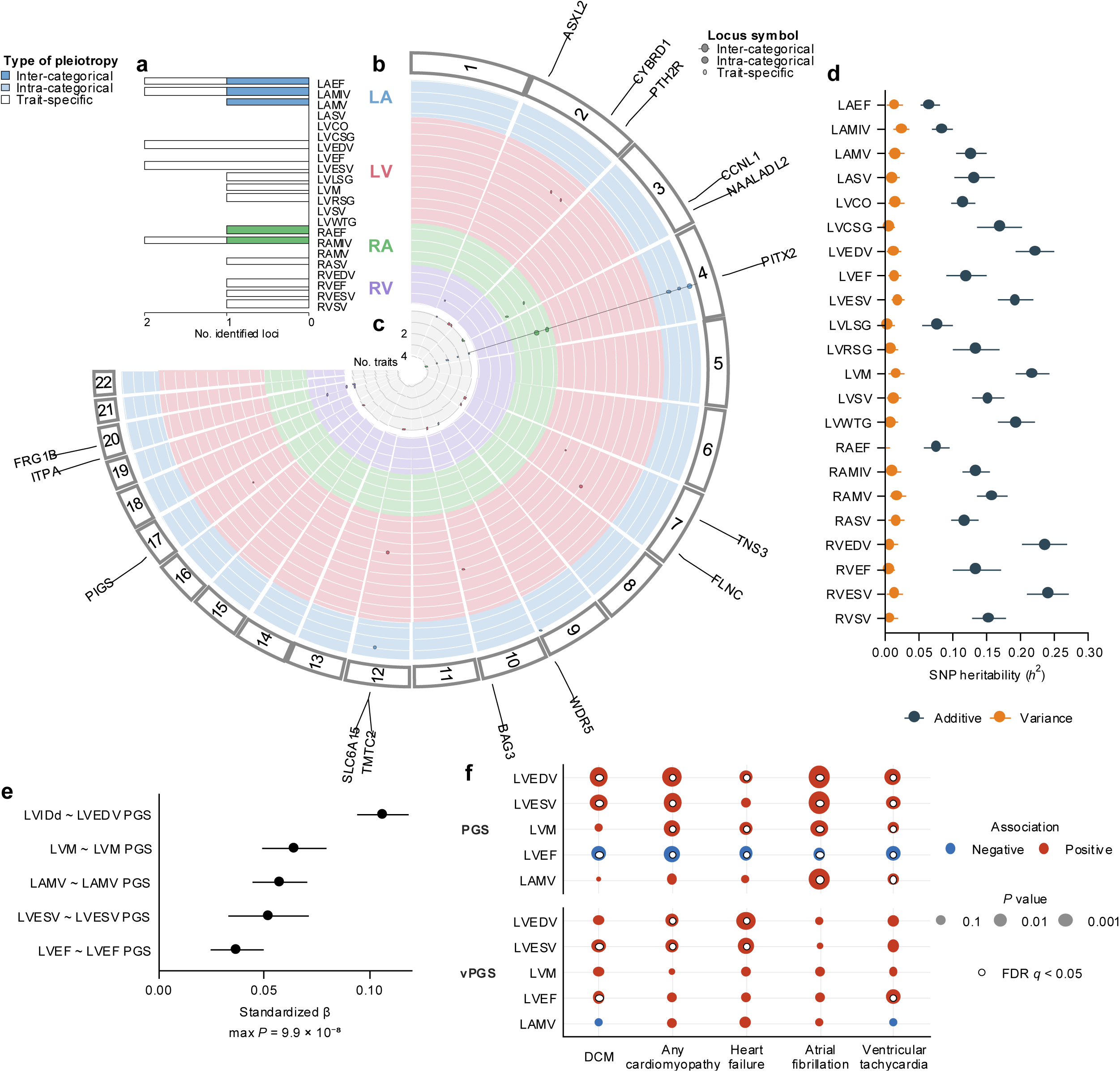
Variance-associated genetics represents a smaller and partly distinct component of cardiac genetic architecture. **a**, Number of genome-wide significant variance quantitative trait loci (vQTLs) identified for each of the 22 CMR phenotypes, classified according to sharing across cardiac chambers. Colors denote LA, LV, RA and RV phenotypes. **b**, Circular representation of genome-wide significant vQTL associations across the cardiac phenotypes. Nearest genes are labeled for inter-categorical loci. **c**, Concentric scale indicating the number of phenotypes associated with each variance-associated region. **d**, SNP-based heritability estimates for additive and variance effects across the 22 CMR phenotypes; points indicate heritability estimates and horizontal bars indicate 95% confidence intervals. **e**, Associations of additive CMR-derived polygenic scores (PGSs) with corresponding echocardiographic phenotypes in Penn Medicine Biobank: LVEDV PGS with LVIDd (*n* = 22,208), LVESV PGS with LVESV (*n* = 9,561), LVM PGS with LVM (*n* = 13,329), LAMV PGS with LAMV (*n* = 18,919) and LVEF PGS with LVEF (*n* = 22,768). Points indicate standardized regression coefficients and horizontal bars indicate 95% confidence intervals from linear regression models adjusted for age and sex. **f**, Associations of additive PGSs and variance polygenic scores (vPGSs) with cardiovascular diagnoses in 55,009 Penn Medicine Biobank participants (DCM, 1,516 cases; any cardiomyopathy, 8,632; heart failure, 12,835; atrial fibrillation, 13,103; ventricular tachycardia, 5,705). Circle size indicates association P value and color indicates association direction (red, positive; blue, negative); white-centered circles indicate FDR q < 0.05. Associations were tested using logistic regression with the corresponding additive PGS and vPGS entered simultaneously, adjusting for enrollment age and sex. Benjamini–Hochberg correction was applied separately across the 25 additive-PGS and 25 vPGS associations.

All 19 variance-associated loci yielded a single credible set by single-trait fine-mapping (Supplementary Table 18), and integrated gene prioritization identified high-confidence candidate genes at 13 credible sets, corresponding to 9 unique genes. *PITX2* was recurrently prioritized across five traits, while additional prioritized genes included *SLC6A15*, *NAALADL2*, *ASXL2* and *RXRA* (Supplementary Table 17).

Genome-wide genetic effects on phenotypic variability were substantially smaller than additive genetic effects on trait levels, with median SNP heritability of 1.36% for variance effects compared with 13.5% for additive effects (Fig. 5d; Supplementary Table 19). Additive and variance heritability were uncorrelated (Pearson r = 0.11, *P* = 0.63).

Consistent with the lower variance heritability, stratified LD score regression showed a more limited and distinct annotation pattern than for additive effects. Among 11 FDR-significant GTEx trait–tissue enrichments, 9 involved nervous-system annotations and were concentrated in right-ventricular phenotypes, with seven annotations enriched for RVEF and two for RVEDV (Supplementary Fig. 5 and Supplementary Table 20). HeartMap analysis identified only two FDR-significant variance enrichments, involving fibroblasts for RVEF and ventricular cardiomyocytes for LVSV (Supplementary Fig. 6 and Supplementary Table 21). Developmental cardiac enhancer analysis identified one FDR-significant variance enrichment, involving LVWTG in a CS17 EnhA1 annotation (Supplementary Fig. 7 and Supplementary Table 22). Together, these findings identified phenotypic variability as a smaller and partly distinct component of cardiac genetic architecture.

### CMR-derived genetic effects generalize to echocardiographic phenotypes and cardiovascular disease

We evaluated whether genetic effects identified from the UK Biobank CMR analyses generalized across cohorts and imaging modalities in Penn Medicine Biobank (PMBB)^31^. Additive polygenic scores showed consistent associations with corresponding echocardiographic phenotypes. The LVEDV polygenic score was associated with left ventricular internal diameter in diastole (LVIDd) (standardized *β* = 0.107, 95% CI 0.094–0.119, *P* = 5.6 × 10^−66^), the LVESV polygenic score with LV systolic volume (*β* = 0.052, 95% CI 0.033–0.071, *P* = 9.9 × 10^−8^), the LVM polygenic score with LV mass (*β* = 0.065, 95% CI 0.049–0.080, *P* = 1.3 × 10^−16^), the LAMV polygenic score with LA maximum volume (*β* = 0.057, 95% CI 0.044–0.071, *P* = 2.3 × 10^−17^), and the LVEF polygenic score with echocardiographic LVEF (*β* = 0.037, 95% CI 0.024–0.050, *P* = 1.2 × 10^−8^) (Fig. 5e; Supplementary Table 23).

Additive CMR-derived polygenic scores were also associated with related cardiovascular diagnoses in PMBB, with 20 of 25 tested associations remaining significant after FDR correction. For example, a higher LVEDV polygenic score was associated with dilated cardiomyopathy (OR = 1.15, 95% CI 1.09–1.21, *P* = 2.2 × 10^−7^), any cardiomyopathy (OR = 1.07, 95% CI 1.04–1.09, *P* = 6.9 × 10^−8^), heart failure (OR = 1.03, 95% CI 1.01–1.05, *P* = 0.0047), atrial fibrillation (OR = 1.10, 95% CI 1.08–1.13, *P* = 9.0 × 10^−19^) and ventricular tachycardia (OR = 1.06, 95% CI 1.03–1.09, *P* = 1.2 × 10^−4^). Conversely, a higher LVEF polygenic score was associated with lower odds of dilated cardiomyopathy (OR = 0.91, 95% CI 0.86–0.96, *P* = 3.9 × 10^−4^), heart failure (OR = 0.97, 95% CI 0.95–0.99, *P* = 0.0015) and ventricular tachycardia (OR = 0.95, 95% CI 0.93–0.98, *P* = 6.2 × 10^−4^). Variance polygenic scores showed more limited disease associations, with 7 of 25 tests remaining significant after FDR correction, including LVEDV vPGS with heart failure (OR = 1.06, 95% CI 1.03–1.08, *P* = 5.5 × 10^−7^), LVESV vPGS with heart failure (OR = 1.05, 95% CI 1.02–1.07, *P* = 5.9 × 10^−5^) and dilated cardiomyopathy (OR = 1.08, 95% CI 1.03–1.14, *P* = 0.0042), and LVEF vPGS with ventricular tachycardia (OR = 1.05, 95% CI 1.02–1.08, *P* = 0.0011) (Fig. 5f; Supplementary Table 24).

Additive and variance polygenic scores were modestly correlated across all 22 CMR traits in PMBB (Pearson *r* = −0.159 to 0.327; N = 59,687), suggesting that they capture partly distinct genetic information. We then tested whether variance polygenic scores generalized specifically to interindividual variability in corresponding echocardiographic phenotypes using QUAIL. Variance polygenic scores for LVEF and LVESV were associated with variability in minimum observed LVEF (*β* = 0.324, 95% CI 0.080–0.569, *P* = 0.0093) and maximum LV systolic volume (*β* = 1.54, 95% CI 0.34–2.74, *P* = 0.012), respectively, after adjustment for the corresponding additive polygenic score. No corresponding associations were observed for the other three traits (Supplementary Table 25).

We also tested whether individual vQTLs replicated in clinically acquired echocardiographic phenotypes. Among six genome-wide significant lead variants with corresponding phenotypes available in PMBB, three were significant after Bonferroni correction in participants of European genetic ancestry: 4:111677041:T:A (rs2634074) for LAMV with left atrial volume (*β* = 0.0155, 95% CI 0.0068–0.0243, *P* = 5.0 × 10^−4^), 7:128469760:G:A (rs3807133) for LVESV with LV systolic volume (*β* = −0.0381, 95% CI −0.0644 to −0.0119, *P* = 0.0044), and 2:172414462:G:A (rs528) for LVEDV with LVIDd as a proxy phenotype (*β* = −2.02 × 10^−4^, 95% CI −3.49 × 10^−4^ to −5.61 × 10^−5^, *P* = 0.0067; Supplementary Table 26). Notably, rs528, a 3′-UTR variant in *CYBRD1*, was associated with *CYBRD1* expression in atrial appendage tissue in GTEx v8 (normalized effect size = −0.097, *P* = 7.1 × 10^−5^)^19^. The rs528 variance association was not accompanied by a significant additive effect in PMBB and occurred at a UKB vQTL lacking a nearby additive locus or evidence of additive–variance colocalization (Supplementary Table 27), consistent with a variance-specific genetic effect. Together, these findings demonstrate that variance-associated genetic effects can generalize across cohorts and imaging modalities at both individual loci and the polygenic level.

## Discussion

We characterized common genetic effects on cardiac structure and function across 22 CMR phenotypes and found extensive genetic sharing across the heart alongside marked heterogeneity among independent signals within individual loci. These signals converged on cardiac developmental and disease biology, while variance mapping identified a smaller, partly distinct component influencing phenotypic variability. Together, these findings distinguish broadly shared cardiac genetic effects from signal-level anatomical specificity and extend the genetic study of cardiac phenotypes beyond additive genetic effects.

Genetic sharing across the heart was particularly apparent at the level of individual fine-mapped signals. Although prior studies have demonstrated substantial genetic correlation and locus-level pleiotropy among cardiac imaging traits^2–4^, shared loci can contain multiple independent genetic effects and therefore do not necessarily imply a common underlying signal. In our multivariate analysis, nearly all fine-mapped signals showed effects across multiple phenotypes and 96% across multiple chambers, with approximately half spanning all four chambers. This sharing nevertheless showed biological organization. Corresponding ventricular measures were particularly strongly connected, and credible-set sharing was greater within atrial or ventricular compartments than between them. Independently, community detection separated phenotypes into groups corresponding broadly to atrial function, chamber remodeling and ventricular systolic function. These patterns were not captured by simple left–right laterality or individual chamber identity, suggesting that common genetic effects follow coordinated aspects of cardiac physiology rather than anatomical boundaries alone. At the same time, *RBM20* and the *TTN*–*CCDC141* region demonstrated that independent signals within the same genomic region can have markedly different phenotypic profiles. This pattern was particularly striking at *RBM20*. The ventricular-patterned single-variant signal was led by rs189569984, a missense variant previously associated with LVESV, RVESV and LVEF^12^, whereas the independent predominantly atrial signal was led by rs10749053, a previously identified atrial-fibrillation susceptibility variant^13^. Thus, distinct signals at the same locus showed previously reported cardiac associations concordant with the anatomical profiles resolved by multivariate fine-mapping. This signal-level differentiation parallels phenotypic heterogeneity reported for rare RBM20 variants, where different amino-acid substitutions at the same position in the protein have been associated with DCM with or without left ventricular noncompaction and with distinct cellular phenotypes^32^. A cardiac locus therefore cannot necessarily be classified as simply pleiotropic or trait-specific; allelic heterogeneity can produce distinct patterns of phenotypic sharing within the same genomic region. This signal-level heterogeneity complements prior evidence that shared genetic pathways can produce divergent phenotypic effects across cardiac traits and cardiomyopathies^33^.

Our findings also connect common genetic variation in adult cardiac structure and function to both cardiac development and mature cardiomyocyte biology. Genes prioritized from fine-mapped associations were enriched for developmental processes including heart and ventricular morphogenesis. Genome-wide heritability enrichment localized predominantly to cardiac tissues and atrial and ventricular cardiomyocytes. These findings were complemented by genome-wide heritability enrichment in regulatory elements active during human cardiac development. Significant enrichment was observed for 16 of 22 CMR traits, spanning all nine assayed Carnegie stages and all three active enhancer states. Notably, LVEDV and LVESV each showed significant enrichment in 51 of 54 developmental enhancer annotations, suggesting particularly widespread overlap between genetic determinants of adult ventricular dimensions and regulatory programs active during cardiac organogenesis. These findings extend prior CMR genetic studies implicating cardiac developmental pathways and congenital heart disease genes in variation in adult cardiac structure and function^2,4^. Together, these observations suggest that common variation influencing adult cardiac phenotypes acts in part through developmental programs that establish cardiac structure, while enrichment in mature cardiomyocytes points to additional effects on the cellular processes that maintain cardiac form and function after development.

Our findings also connect genetic determinants of quantitative cardiac phenotypes with susceptibility to cardiomyopathy and heart failure. Prior studies have linked common genetic variation in CMR-derived ventricular traits to cardiomyopathy and heart failure through polygenic prediction, genetic correlation and shared genetic signals^2–4,34^. CMR-prioritized genes were enriched for established cardiomyopathy genes, and a subset of CMR-associated regions contained shared genetic signals with HCM, DCM or heart failure. This extends prior work showing that genetic analyses of quantitative cardiac endophenotypes in largely disease-free populations can recover genes and pathways implicated in clinical cardiovascular disease^34^. At *TMEM182*, the ventricular-patterned single-variant signal was itself a previously identified HCM lead variant, while prior LV eQTL colocalization implicated lower LV *TMEM182* expression in DCM risk^23,28^. At *FHOD3*, independent intronic and coding signals showed different patterns of quantitative cardiac involvement; the intronic signal was led by a variant in strong LD with a prior HCM lead and colocalized with DCM in our analysis, whereas the coding p.Val822Phe signal was previously fine-mapped to resting heart rate^29^. Together with the distinct signals at *RBM20*, these findings show that different variants within the same cardiac locus can map to different cardiac phenotypes and anatomical patterns. At *BAG3*, the lead variant of a broad CMR signal colocalizing with HCM, DCM and heart failure was in near-perfect LD with the *BAG3* missense variant p.Cys151Arg (rs2234962). p.Cys151Arg has opposing associations with cardiomyopathy, being associated with lower DCM risk but higher HCM risk, and has also been associated with lower DCM risk among carriers of *TTN* truncating variants in highly expressed exons^27^. Together, these findings support partial continuity between common genetic effects underlying quantitative cardiac variation and those contributing to pathological cardiac remodeling.

A second distinct feature of the study was the identification of common genetic effects on phenotypic variability rather than additive trait effects. Variance QTLs can prioritize loci at which genetic effects may depend on genetic or environmental context, providing information complementary to conventional mean-based association testing^8^. Variance-associated heritability was substantially lower than additive heritability, and most vQTLs lacked a nearby genome-wide significant additive association, indicating that variance mapping captures information not readily recovered from conventional GWAS. The recurrent prioritization of *PITX2* across additive and variance analyses implicates the same locus in genetic effects on both phenotype level and variability, although the underlying mechanisms remain uncertain. Importantly, variance effects showed evidence of generalization beyond UK Biobank: variance polygenic scores for LVEF and LVESV were associated with variability in corresponding clinical echocardiographic measurements, and several individual vQTLs replicated across cohorts and imaging modalities. These findings identify phenotypic variability as a smaller and partly distinct component of cardiac genetics. However, vQTL association alone does not identify the genetic or environmental factor responsible for the variance effect, and the relevant modifying contexts remain to be determined^8^.

Several limitations should be considered. First, the UK Biobank imaging cohort is predominantly of European genetic ancestry and exhibits healthy-volunteer selection relative to the broader UK Biobank cohort, which may limit generalizability^35^. Second, multivariate fine-mapping depends on model assumptions regarding the number and sharing of genetic effects, and the extensive cross-phenotype support observed here should therefore be interpreted in the context of those assumptions^6^. Third, HyPrColoc assumes at most one causal variant for each trait within a region, and colocalization supports a shared genetic signal but does not by itself establish the underlying biological mechanism^26^. Fourth, vQTL mapping identifies loci associated with phenotypic variance but does not identify the genetic or environmental context responsible for that variance^8^. Finally, PMBB provided an independent clinically ascertained cohort, but echocardiographic phenotypes were not always direct analogues of the CMR measurements and only a subset of vQTLs could be evaluated. Nevertheless, convergence across fine-mapping, functional annotation, disease genetics and independent clinical phenotypes supports the principal patterns observed.

In summary, common genetic effects on cardiac structure and function are broadly shared across the heart, yet individual loci can contain independent signals with distinct anatomical and phenotypic profiles. Variance-associated genetics adds a smaller, partly distinct dimension, extending the genetic study of cardiac phenotypes beyond additive genetic effects.

## Methods

### Study population and relatedness

We analyzed cardiac magnetic resonance imaging-derived phenotypes from UK Biobank Category 157, generated using the automated image-segmentation pipeline developed by Bai et al. (2020)^9^. The discovery analyses included up to 77,271 participants and up to 10,662,572 variants. Participants were restricted to an unrelated analysis set using UK Biobank KING kinship estimates^36^. Pairs with kinship coefficients greater than 0.0884 were represented as edges in a relatedness graph. All isolated participants were retained, and a maximum independent vertex set was selected within each connected component using igraph::largest_ivs, thereby retaining the largest possible unrelated subset within each component.

### Cardiac phenotypes and covariates

We analyzed a panel of 22 cardiac imaging traits. These comprised left atrial ejection fraction (LAEF), minimum volume (LAMIV), maximum volume (LAMV) and stroke volume (LASV); left ventricular ejection fraction (LVEF), end-diastolic volume (LVEDV), end-systolic volume (LVESV), stroke volume (LVSV), cardiac output (LVCO), myocardial mass (LVM), global mean myocardial wall thickness (LVWTG), global longitudinal strain (LVLSG), global circumferential strain (LVCSG) and global radial strain (LVRSG); right atrial ejection fraction (RAEF), minimum volume (RAMIV), maximum volume (RAMV) and stroke volume (RASV); and right ventricular end-diastolic volume (RVEDV), ejection fraction (RVEF), end-systolic volume (RVESV) and stroke volume (RVSV).

Each trait was analyzed among participants with complete phenotype and covariate data. Covariates comprised age at recruitment, genetic sex and the first five genetic principal components.

### Genotype data preparation

UK Biobank imputed BGEN v3 genotype data for autosomes 1–22 were converted to chromosome-specific PLINK files for the retained participant set. BGEN alleles were interpreted using the reference-first convention, and variants with minor allele frequency below 0.005 were excluded.

### Genome-wide additive association analysis

Genome-wide additive association analysis was performed separately for each of the 22 CMR phenotypes using PLINK2 linear regression^37^. For each trait, the original CMR phenotype was regressed on genotype under an additive genetic model, adjusting for age at recruitment, genetic sex and the first five genetic principal components.

### Summary-statistic quality control and coordinate conversion

Summary statistics were matched to UK Biobank MFI records by chromosome position and unordered allele pair. MFI records with conflicting duplicate annotations were excluded. Variants were required to have nonmissing INFO, a minor allele frequency between 0 and 0.5 and an MFI minor allele matching one summary-statistic allele. We retained variants with INFO greater than 0.8 and minor allele count greater than 20, where minor allele count was calculated as 2N × MAF. GRCh38 coordinates were generated with GWASLab v4.1.4, retaining unique mappings that remained on the same chromosome.

### Locus definition and cross-locus merging

Loci were defined separately for additive and variance statistics using 50-kb genomic tiles. Tiles containing at least one variant with *P* ≤ 1 × 10^−5^ were marked as occupied, directly adjacent occupied tiles were merged and each resulting interval was padded by one 50-kb tile on either side. A padded interval was retained when it contained at least one genome-wide significant variant with *P* ≤ 5 × 10^−8^; the most significant variant in the interval was designated the lead variant. Within each trait, association model and chromosome, intervals were merged when they overlapped or when their lead variants had Pan-UKB European ancestry linkage disequilibrium^38^ *r*^2^ > 0.001. Merging was applied transitively through connected components, with final boundaries defined by the minimum start and maximum end. Pan-UKB variants were matched by GRCh37 position and unordered allele pair, requiring a unique reference match. Variance loci were annotated by the nearest genome-wide significant additive variant and whether it was located within 500 kb.

### Locus coordinate conversion

Final locus boundaries and lead-variant positions were converted from GRCh37 to GRCh38 using rtracklayer^39^ with an hg19-to-hg38 chain file. Lower boundary, upper boundary and lead position were first lifted as separate single-base coordinates; the conversion was accepted when all three mapped exactly once to the same chromosome. When this criterion failed but the lead position mapped uniquely, the complete GRCh37 interval was lifted as a fallback. Mapped fragments were restricted to the chromosome containing the lifted lead variant, and GRCh38 boundaries were defined by the outermost retained fragment coordinates. Converted loci were retained only when both boundaries and the lead position were valid and chromosome-consistent.

### Comparison with previous cardiac imaging GWAS

Additive loci were compared with genome-wide significant variants from three previous cardiac imaging GWAS^2–4^. Variants with *P* ≤ 5 × 10^−8^ were retained from each study. For each locus, prior variants were classified as directly overlapping the interval, within 500 kb of an interval edge, between 500 kb and 2 Mb or more than 2 Mb away. Comparisons were conducted using traits present in the Pirruccello 2020, Pirruccello 2022 and Nicholls 2026 datasets, both within the same trait and across any mapped trait, comprising LAEF, LAMV, LAMIV, LASV, LVEF, LVEDV, LVESV, LVSV, LVM, RVEF, RVEDV, RVESV, RVSV. A locus was classified as novel relative to the included studies only when every expected comparison was evaluable and no previously reported genome-wide significant variant was located within 2 Mb of either locus boundary.

### Multivariate fine-mapping

Multivariate fine-mapping was performed for an 18-trait cardiac panel comprising LAEF, LAMIV, LAMV, LVCO, LVCSG, LVEDV, LVEF, LVESV, LVLSG, LVM, LVRSG, LVWTG, RAEF, RAMIV, RAMV, RVEDV, RVEF and RVESV^6^. After finding substantial linear dependence and numerical instability in the multivariate correlation matrices, the four stroke-volume traits (LASV, LVSV, RASV and RVSV) were excluded because they are directly derived from the corresponding volumetric measures. Unique additive loci from the 22-trait source panel were merged transitively when their intervals overlapped within a chromosome. One region overlapping the major histocompatibility complex was excluded from multivariate fine-mapping because a stable fine-mapping solution could not be obtained. Regions were required to contain complete (*Z*) statistics across all 18 traits. European-ancestry linkage disequilibrium matrices were used for each region. A residual-correlation matrix was estimated from two putatively null variants per region, selected as the variants with the smallest maximum absolute *Z* statistic across the 18 traits among variants with maximum *Z* < 2. The uncentered second moment of the selected *Z*-score vectors was calculated and scaled to a correlation matrix.

An empirical covariance prior was learned from one lead variant per region, defined as the complete variant with the largest maximum absolute (*Z*) statistic. Regions were included in prior training when the selected variant reached *P* ≤ 5 × 10^−8^ in at least one trait. Candidate covariance matrices included an empirical second-moment covariance, FLASH-derived factors, principal-component covariances and canonical covariance matrices, which were combined using Bovy extreme deconvolution^40^. For each region, trait-specific sample size was defined as the median finite per-variant (*N*), and regional sample size as the median across traits.

mvSuSiE was fit with *L* = 10, 95% credible-set coverage, minimum absolute credible-set correlation of 0.5 and a trait-support local false sign rate threshold of 0.01. Prior variance was estimated by expectation-maximization, whereas residual variance and prior mixture weights were fixed.

### Bipartite network analysis of cross-trait genetic sharing

Cross-trait genetic sharing was represented as a binary bipartite network linking mvSuSiE credible sets to supported cardiac phenotypes. Rows of the incidence matrix represented credible sets and columns represented the 18 analyzed phenotypes. An edge connected a credible set to a phenotype when the corresponding single-effect local false sign rate was ≤0.01 and the conditional effect was finite and nonzero.

Community structure was identified directly from the bipartite network by maximizing Barber bipartite modularity^11^. The observed network was analyzed using 100 optimization iterations, and phenotype assignments from the maximum-modularity solution were retained. Barber bipartite modularity is denoted Q.

To determine whether the observed modular organization exceeded that expected from differences in node connectivity alone, we generated 10,000 randomized binary networks preserving both row and column degree sequences. Thus, each randomized network retained the number of phenotypes supported by each credible set, the number of credible sets supporting each phenotype and the total number of edges, while randomizing the identities of individual connections. Preservation of both degree sequences and the total edge count was verified for every randomized network, and community detection was repeated using the same procedure. The one-sided empirical P value was calculated as 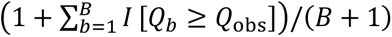, where *B* = 10,000.

Robustness of the phenotype partition to arbitrary node ordering was evaluated by independently permuting the credible-set rows and phenotype columns of the unchanged observed incidence matrix 10,000 times. Community detection was repeated after each permutation, and phenotype assignments were mapped back to their original identities. Agreement with the reference partition was quantified by the proportion of exact partition recoveries and the adjusted Rand index.

### Anatomical organization of cross-trait genetic sharing

To test whether genetic sharing followed prespecified cardiac anatomy, pairwise credible-set sharing was compared according to atrial versus ventricular compartment, left versus right laterality and individual cardiac chamber. For each phenotype pair, normalized sharing was calculated as the number of credible sets supporting both phenotypes divided by the number supporting either phenotype, corresponding to the Jaccard index *J_ij_* = |*C_i_* ∩ *C_j_*|/|*C_i_* ∪ *C_j_*|.

For each anatomical classification, mean normalized sharing among phenotype pairs assigned to the same group was compared with that among phenotype pairs assigned to different groups. Statistical significance was evaluated using 10,000 permutations of the anatomical labels across phenotype nodes while preserving the observed credible-set sharing network and the number of phenotypes assigned to each group. One-sided empirical *P* values were calculated as 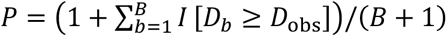, where *D* denotes the difference between mean within-group and between-group sharing and *B* = 10,000.

### Functional annotation of multivariate credible-set variants

Unique GRCh37 variants from mvSuSiE credible sets were submitted in VCF-like representation to the Ensembl GRCh37 regional Variant Effect Predictor^41^ REST endpoint. Transcript annotations were restricted to official MANE Select release 1.5.^42^ When more than one MANE Select annotation remained for a variant, annotations were ranked by VEP consequence severity, impact, gene symbol and transcript identifier. Coding, splice and VEP HIGH, MODERATE or LOW impact annotations were retained for downstream displays.

### Single-trait fine-mapping

Additive and variance loci were fine-mapped with SuSiE-RSS^43^ using the Pan-UKB European ancestry linkage disequilibrium reference panel. Variants were required to have finite effect estimates, standard errors and *P* values, with standard errors greater than zero; *Z* statistics were calculated as *β*/SE. Alleles were aligned to the Pan-UKB reference. Loci with fewer than 20 variants after harmonization were excluded. The effective sample size was the median per-variant *N* within each harmonized locus. SuSiE-RSS was fit with a maximum of *L* = 10 effects, 99% credible-set coverage, minimum absolute credible-set correlation of 0.5 and fixed residual variance.

### Gene-level association and PoPS prioritization

Gene-level association testing was performed with MAGMA v1.07a^44^ using the snp-wise=mean model, GRCh38 1000 Genomes^45^ reference genotypes and per-variant SNP identifiers, *P* values and sample sizes. PoPS gene prioritization^46^ was performed from the MAGMA gene results using the supplied full feature collection and control-feature matrix. MAGMA gene-property analyses used GTEx v8 tissue-expression covariates.

### Integrated gene prioritization

FLAMES^15^ was used to integrate single-trait fine-mapping, MAGMA gene-level associations, PoPS scores and GTEx tissue-property results. Annotation and scoring were performed separately for each cardiac trait and association model (additive and variance).

### Gene Ontology biological-process enrichment

Biological-process enrichment was evaluated among high-confidence genes prioritized by FLAMES from additive analyses. Genes with estimated cumulative precision greater than 0.75 were pooled across the 22 CMR traits and deduplicated by gene symbol. Enrichment analysis was performed using Enrichr and the GO_Biological_Process_2025 database^16,17^. Gene Ontology terms with Benjamini-Hochberg-adjusted *P* < 0.05 were considered significantly enriched.

### Enrichment of curated cardiac disease genes

Additive FLAMES-prioritized genes were tested for overrepresentation of curated hypertrophic cardiomyopathy^7^, dilated cardiomyopathy^7^ and arrhythmogenic cardiomyopathy gene sets^22^. Ensembl gene-version suffixes were removed before matching genes by Ensembl gene ID. The gene universe comprised unique genes in the additive FLAMES raw table, and the selected set comprised unique additive FLAMES-predicted genes present in that universe. For each curated gene set, enrichment was tested using a one-sided hypergeometric test, where *N* was the background size, *n* the selected-set size, *K* the number of curated genes in the background and *x* their overlap with the selected set. A one-sided Fisher exact test was used as a cross-check and its odds ratio was reported. Benjamini-Hochberg correction was applied across the three disease-specific tests. The union of the three curated sets was additionally analyzed with shared genes counted once.

### GTEx tissue-specific stratified LD Score Regression

Tissue-specific heritability enrichment was evaluated using GTEx^19^ entries from the LDSC multi-tissue gene-expression annotation manifest. Stratified LD Score Regression^18^ incorporated baselineLD v2.2^47^, HapMap3 weights excluding the major histocompatibility complex, European frequency files and overlapping annotations. Additive and variance-association summary statistics were analyzed separately. Benjamini-Hochberg correction was applied across GTEx tissue annotations within each trait and association model.

### Cardiac cell-type annotations

Cardiac cell-type annotations were constructed from HeartMap^20^ global-AUC marker genes. One gene set was defined for each of 14 HeartMap cell types together with an all-genes control set. Marker genes were mapped through Ensembl gene identifiers, and binary annotations were created from 100-kb windows around selected genes. Annotation LD scores were calculated using 1000 Genomes Phase 3 European genotypes, a 1-cM window and thin-annotation format.

### Cardiac cell-type stratified LD Score Regression

Cardiac trait summary statistics were munged against the HapMap3 SNP list while retaining per-variant sample sizes. Cell-type-specific LD Score Regression was performed for 22 cardiac traits across the 14 HeartMap annotations using baselineLD v2.2, HapMap3 weights excluding the major histocompatibility complex, European frequency files and overlapping-annotation calculations. Additive and variance-association summary statistics were analyzed separately. Benjamini-Hochberg correction was applied across the 14 cell-type annotations within each trait and association model.

### Developmental cardiac enhancer enrichment

Developmental cardiac enhancer annotations were derived from Heart Hub hg19 ChromHMM 25-state data^21^. The enhancer states 13_EnhA1, 14_EnhA2 and 15_EnhAF were retained across 18 cardiac developmental samples, yielding 54 sample-by-state annotations. Intervals were converted to autosomal BED annotations and then to binary SNP annotations. Annotation LD scores were computed using 1000 Genomes Phase 3 European genotypes, a 1-cM window and thin-annotation format. Each annotation was fit jointly with baselineLD v2.2 using HapMap3 regression variants excluding the major histocompatibility complex, European frequency files and overlapping-annotation coefficients. Enrichment was assessed from the positive coefficient tail, and Benjamini-Hochberg correction was applied across the 54 annotations within each trait. Additive and variance-association summary statistics were analyzed separately for each trait.

### Cross-trait colocalization with cardiomyopathy and heart failure

Cross-trait colocalization was evaluated between the 22 additive CMR traits and hypertrophic cardiomyopathy^23^, dilated cardiomyopathy^24^ and heart failure^25^ using HyPrColoc^26^. Additive loci were merged by overlapping GRCh37 intervals. Within each region, summary statistics for all 25 traits were matched by GRCh37 chromosome position and unordered allele pair, and effect estimates were aligned to a common allele. Variants were required to have finite effect estimates and standard errors greater than zero and to be present with concordant alleles across all 25 traits; regions with fewer than 10 shared variants were excluded. HyPrColoc was applied using the default priors (*P*_1_ = 1 × 10^−4^, *P*_c_ = 0.02) and regional association and alignment probability thresholds of 0.5, without an external linkage-disequilibrium matrix. Trait clusters, candidate variants and colocalization posterior probabilities were retained. For highlighted comparisons with previously reported disease-associated variants, pairwise linkage disequilibrium was calculated from the corresponding Pan-UKB European-ancestry regional LD matrix used for multivariate fine-mapping.

### Genome-wide variance QTL association analysis

Genome-wide variance QTL analyses were performed separately for each of the 22 CMR phenotypes using QUAIL^8^. For each trait, QUAIL Step 1 was applied among participants with complete phenotype and covariate data across 2,000 quantile levels to construct a quantile-integrated rank-score phenotype. Models included age at recruitment, genetic sex and the first five genetic principal components, and trait datasets were required to contain unique participant identifiers and at least 50 complete observations.

Genome-wide variance association testing was then performed in QUAIL Step 2. For each variant, PLINK2 linear regression was used to test the association between genotype and the trait-specific quantile-integrated rank score, adjusting for the same covariates. Variance effect estimates, standard errors, test statistics, *P* values and per-variant sample sizes were retained for downstream analyses.

### SNP heritability and genetic correlation

LD Score Regression^48^ was used to estimate SNP heritability separately for additive and variance association statistics, and cross-trait LD Score Regression^10^ was used to estimate pairwise genetic correlations from additive association statistics. Across the 22 phenotypes, the relationship between additive and variance SNP heritability estimates was assessed using Pearson correlation.

### Polygenic-score weight estimation

Additive and variance polygenic-score weights were estimated separately with PRS-CSx^49^ from GRCh37 summary statistics. Variants were restricted to HapMap3 rsIDs represented in the UK Biobank European linkage disequilibrium reference and were supplied with SNP identifier, effect allele, other allele, effect estimate and standard error. PRS-CSx was run separately by chromosome with European target population, chromosome-specific median per-variant sample size, automatic global shrinkage (*φ* = auto) and prior parameter *b* = 0.5.

### Polygenic-score calculation in the Penn Medicine Biobank

Additive and variance polygenic scores for all 22 CMR traits were calculated for Penn Medicine Biobank^31^ participants with available genotypes (release 2024-3.0) using pgsc_calc v2.2.0^50^. The pgsc_HGDP+1kGP_v1 ancestry reference was used, a minimum score–target variant overlap of 70% was required and the ancestry-normalized *Z*_norm2_ value was used in downstream analyses. Pearson correlations between matched additive and variance polygenic scores were calculated across all 22 traits in 59,687 PMBB participants.

### Echocardiographic phenotype processing in PMBB

Echocardiographic measurements were harmonized from CUPID and ProSolv extracts. CUPID stress, transesophageal, intracardiac, congenital transesophageal and research echocardiography procedures were excluded, whereas ProSolv records were restricted to complete or limited transthoracic echocardiograms. Participant identifier, examination identifier and date, procedure, measurement name, value and unit were standardized across sources. Numeric values were parsed after unit removal, and reported numeric ranges were converted to their midpoint. Trait-specific exact measurement-name mappings and physiological bounds were applied. Within each examination, a prespecified preferred measurement was selected. Participant-level summaries included the median, minimum, maximum, interquartile range and number of measurements, together with dates corresponding to the median-nearest, minimum and maximum values.

### Imaging validation of PGS and vPGS

Additive and variance polygenic scores were evaluated against corresponding echocardiographic phenotypes in Penn Medicine Biobank. Five CMR-to-echocardiography mappings were evaluated: LVEF with LVEF, LVEDV with left ventricular internal diameter in diastole, LVESV with LV systolic volume, LVM with LV mass, and LAMV with LA maximum volume. Additive PGS portability was evaluated using participant-level median echocardiographic measurements. Associations were tested by linear regression of each phenotype on the ancestry-normalized *Z*_norm2_ score, adjusting for enrollment age and sex.

Variance PGS validation was performed for the same five CMR traits using the QUAIL vPGS framework. Participant-level minimum LVEF was used for LVEF and maximum measurements were used for the remaining phenotypes. Models used 2,000 quantile levels and included enrollment age, sex and the corresponding additive polygenic score as covariates. Benjamini-Hochberg correction was applied across the five variance-PGS imaging-validation tests.

### Replication of individual variance QTLs

Genome-wide significant vQTL lead variants were tested in PMBB when a corresponding echocardiographic phenotype was available. Six variants were evaluated across four CMR traits: LAMV with left atrial volume, LVESV with LV systolic volume, LVM with LV mass, and LVEDV with LVIDd. For each phenotype, QUAIL Step 1 was applied across 2,000 quantile levels with age, genetic sex, the first six genetic principal components and sequencing batch as covariates. Lead variants were then tested for association with the resulting QUAIL variance phenotype by linear regression of imputed effect-allele dosage, adjusting for the same covariates. Analyses were performed in participants of European genetic ancestry. Statistical significance was defined using Bonferroni correction for six prespecified tests (*P* < 0.0083), with replication additionally requiring the same direction of effect as in UK Biobank.

To assess colocalization between variance-and additive-associated signals in UK Biobank, overlapping vQTL loci were merged into variance regions. Within each region, HyPrColoc was applied jointly to variance-effect estimates for the variance-associated trait(s) and additive-effect estimates across all 22 CMR phenotypes using variants shared across the included traits. Clusters containing at least one variance and one additive trait were retained as mixed additive–variance colocalization signals.

### Cardiovascular disease associations of polygenic scores

Additive and variance polygenic scores for LVEF, LVM, LVEDV, LVESV and LAMV were tested for association with cardiovascular diagnoses in the Penn Medicine Biobank. Diagnoses were defined from PheCodeX^51^ records as dilated cardiomyopathy (CV_414.2), any cardiomyopathy (CV_414), heart failure (CV_424), atrial fibrillation (CV_416.21) and ventricular tachycardia (CV_416.12). Scores were standardized within the analysis sample, and the corresponding additive and variance scores were included simultaneously in logistic regression models adjusting for enrollment age and sex. Benjamini-Hochberg correction was applied separately across the 25 additive-score and 25 variance-score associations.

### Software

Analyses were conducted on the UK Biobank Research Analysis Platform (DNAnexus) and the Penn Medicine LPC cluster. Relatedness graph analyses were performed using igraph. Additive quantitative trait locus analyses were performed using PLINK v2.00a3LM. Variance quantitative trait locus analyses were performed with QUAIL (commit f582eaf). Summary-statistic harmonization and coordinate conversion used GWASLab v4.1.4. Multivariate fine-mapping was performed in R v4.5.2 using mvsusieR v0.3.0, susieR v0.16.5, mashr v0.2.79 and flashier v1.0.7. Cross-trait colocalization was performed with HyPrColoc v1.0. Bipartite community analysis was performed using BipartiteModularityMaximization v1.23.120.1, with degree-preserving network randomization performed using vegan v2.7.2. Single-trait fine-mapping was performed in R v4.5.2 using susieR v0.16.5. Gene-level association and prioritization analyses used MAGMA v1.07a, PoPS (commit 76eb86c) and FLAMES (commit d411689). SNP heritability and genetic correlation analyses used ldscr v0.2.1, and stratified LD Score Regression used LDSC (commit aa33296). Polygenic-score weights were estimated with PRS-CSx (based on commit 5f27ff8, with Python compatibility modifications), and scores were calculated with pgsc_calc v2.2.0.

## Data availability

Individual-level UK Biobank data analyzed in this study are available to approved researchers through the UK Biobank application process (https://www.ukbiobank.ac.uk/). This study was conducted under UK Biobank application number 194410. Individual-level Penn Medicine Biobank data are subject to institutional access restrictions. GWAS summary statistics generated in this study are available from the corresponding author upon reasonable request, subject to applicable data-sharing requirements.

## Ethics oversight

The North West–Haydock Research Ethics Committee of the NHS Health Research Authority granted ethical approval for UK Biobank as a Research Tissue Bank (REC reference 21/NW/0157). This approval covers qualifying secondary analyses of UK Biobank data without requiring separate project-specific ethical approval. All UK Biobank participants provided informed consent. The present study was conducted under UK Biobank application number 194410.

The Institutional Review Board of the University of Pennsylvania granted ethical approval for the Penn Medicine Biobank analyses under protocol 25-1362.

## Acknowledgements

This research has been conducted using the UK Biobank Resource under application number 194410.

We acknowledge the Penn Medicine BioBank (PMBB) for providing data and thank the patient-participants of Penn Medicine who consented to participate in this research program. We would also like to thank the Penn Medicine BioBank team and Regeneron Genetics Center for providing genetic variant data for analysis. The PMBB is approved under IRB protocol #813913 and supported by the Perelman School of Medicine at the University of Pennsylvania, a gift from the Smilow family, and the National Center for Advancing Translational Sciences of the National Institutes of Health under CTSA award number UL1TR001878.

## Funding

This work was supported in part by the U.S. Department of Veterans Affairs (IK2-BX006551); the National Heart, Lung, and Blood Institute of the National Institutes of Health (R01HL141232, R01HL168841, and R35HL177035); and the National Institute of General Medical Sciences of the National Institutes of Health (R35GM119465).

The content is solely the responsibility of the authors and does not necessarily represent the official views of the National Institutes of Health. This publication does not represent the views of the U.S. Department of Veterans Affairs or the United States Government.

## Competing interests

M.G.L. reports receiving research grants from MyOme and personal fees from BridgeBio. S.M.D. reports receiving consulting fees from Tourmaline Bio and in-kind research support from Amgen and Novo Nordisk, unrelated to the current work.

## Notes

### Author Declarations

The North West-Haydock Research Ethics Committee of the NHS Health Research Authority granted ethical approval for UK Biobank as a Research Tissue Bank (REC reference 21/NW/0157). This approval covers qualifying secondary analyses of UK Biobank data without requiring separate project-specific ethical approval. All UK Biobank participants provided informed consent. The present study was conducted under UK Biobank application number 194410. The Institutional Review Board of the University of Pennsylvania granted ethical approval for the Penn Medicine Biobank analyses under protocol 25-1362.

