## Supplementary Information for "Shared additive- and variance-associated genetic architecture of cardiac structure and function"

Supplementary Tables 1–27 are provided in the accompanying Excel file. Captions are provided below.

### Supplementary Table captions

**Supplementary Table 1. SNP-based heritability estimates for additive CMR associations**. LD Score Regression estimates across 22 CMR-derived phenotypes. h^2^ denotes SNP-based heritability; SE denotes standard error. The table additionally reports the LD Score Regression intercept and associated standard error and *P* value.

**Supplementary Table 2. Genome-wide significant additive locus–trait associations and comparison with previous CMR GWAS**. Results are shown for all 748 genome-wide significant additive locus–trait associations across 22 CMR phenotypes. The table reports locus boundaries and lead variants, cross-phenotype merged-region membership and pleiotropy classification, and comparison with previous CMR GWAS for the 13 phenotypes with corresponding prior data. Novelty is reported both relative to the same phenotype and across all 13 comparable phenotypes. Genomic coordinates are reported in GRCh37.

**Supplementary Table 3. Pairwise genetic correlations among CMR phenotypes**. Cross-trait LD Score Regression estimates for all 231 pairs of the 22 CMR-derived phenotypes. The table reports genetic correlation (r_g_), standard error, *P* value and Benjamini–Hochberg FDR *q* value.

**Supplementary Table 4. Multivariate fine-mapping credible sets across 18 CMR phenotypes**. Summary of the 669 credible sets identified by mvSuSiE. The table reports regional coordinates, credible-set size and purity, sentinel variant and posterior inclusion probability (PIP), and the number and identity of supported phenotypes and cardiac chambers. Phenotype support was defined using a local false sign rate ≤0.01. Genomic coordinates are reported in GRCh37.

**Supplementary Table 5. Phenotypic support for multivariate fine-mapping credible sets**. Trait-specific conditional effects for mvSuSiE credible sets across the 18 phenotypes included in multivariate fine-mapping. Rows are shown for credible-set–phenotype pairs meeting the phenotype-support criterion of local false sign rate ≤0.01 with a finite nonzero conditional effect.

**Supplementary Table 6. Phenotype communities identified from the bipartite credible-set sharing network**. Barber bipartite community assignments for the 18 phenotypes included in multivariate fine-mapping. The table reports the assigned community, descriptive community label and number of credible sets supporting each phenotype.

**Supplementary Table 7. Validation of bipartite community structure**. Summary statistics for the observed Barber bipartite modularity, 10,000 degree-preserving randomized networks and 10,000 random reorderings of the observed incidence matrix. Reported measures include the observed and null modularity distributions, empirical *P* value, exact partition-recovery frequency and adjusted Rand index.

**Supplementary Table 8. Pairwise credible-set sharing across CMR phenotypes**. Pairwise sharing among the 18 phenotypes included in multivariate fine-mapping. The table reports the number of credible sets supporting each phenotype, the intersection and union of supported credible sets, Jaccard similarity and corresponding percentage of shared genetic signals, together with atrial–ventricular, left–right and chamber classifications.

**Supplementary Table 9. Anatomical organization of cross-trait credible-set sharing**. Comparison of mean pairwise Jaccard similarity within versus between prespecified atrial–ventricular, left–right and individual-chamber groupings. Statistical significance was evaluated using 10,000 permutations of anatomical labels across phenotype nodes; one-sided empirical *P* values are reported.

**Supplementary Table 10. Single-trait additive fine-mapping and integrated gene prioritization**. Summary of 650 credible sets identified by SuSiE-RSS from additive CMR associations. The table reports credible-set size, posterior mass, maximum variant PIP and purity, together with high-confidence FLAMES-prioritized genes where available. High-confidence genes were defined by estimated cumulative precision >0.75. Genomic coordinates are reported in GRCh37.

**Supplementary Table 11. Gene Ontology Biological Process enrichment of additive CMR-prioritized genes**. All Gene Ontology Biological Process terms significant after Benjamini–Hochberg correction (FDR <0.05) among the 149 unique high-confidence genes prioritized from additive CMR associations. The table reports enrichment statistics and overlapping genes.

**Supplementary Table 12. GTEx tissue-specific stratified LD Score Regression results for additive CMR associations**. Results are shown for all 53 GTEx tissue annotations across each of the 22 CMR phenotypes. The table reports annotation coefficients, standard errors, *Z* scores and one-sided coefficient *P* values. FDR *q* values were calculated by Benjamini–Hochberg correction across the 53 annotations within each phenotype.

**Supplementary Table 13. HeartMap cardiac cell-type stratified LD Score Regression results for additive CMR associations**. Results are shown for all 14 HeartMap cardiac cell-type annotations across each of the 22 CMR phenotypes. The table reports annotation coefficients, standard errors, *Z* scores and one-sided coefficient *P* values. FDR *q* values were calculated by Benjamini–Hochberg correction across the 14 annotations within each phenotype.

**Supplementary Table 14. Developmental cardiac enhancer stratified LD Score Regression results for additive CMR associations**. Partitioned heritability results are shown for 54 Heart Hub sample-by-state annotations (18 developmental heart samples × EnhA1, EnhA2 and EnhAF) across each of the 22 CMR phenotypes. The table reports annotation composition, heritability enrichment and annotation coefficients. One-sided positive-tail coefficient *P* values were used for Benjamini–Hochberg correction across the 54 annotations within each phenotype.

**Supplementary Table 15. Enrichment of curated cardiomyopathy genes among additive CMR-prioritized genes**. High-confidence FLAMES-prioritized genes were tested for enrichment in curated hypertrophic cardiomyopathy, dilated cardiomyopathy and arrhythmogenic cardiomyopathy gene sets and their union. The table reports observed and expected overlap, fold enrichment, Fisher odds ratios, one-sided hypergeometric *P* values and overlapping genes. Benjamini–Hochberg correction was applied across the three disease-specific tests.

**Supplementary Table 16. High-probability colocalization between CMR traits and cardiomyopathy or heart failure**. Merged additive CMR regions containing at least one HyPrColoc cluster involving hypertrophic cardiomyopathy, dilated cardiomyopathy or heart failure with posterior probability >0.7. The table reports CMR and disease traits within each cluster, posterior probabilities and candidate variants. Genomic coordinates are reported in GRCh37.

**Supplementary Table 17. Genome-wide significant variance QTL associations, fine-mapping and gene prioritization**. Results are shown for all 19 genome-wide significant variance locus–trait associations across 14 CMR phenotypes. The table reports locus and lead-variant information, cross-phenotype sharing, proximity to genome-wide significant additive associations, single-trait fine-mapping results and high-confidence FLAMES-prioritized genes. Genomic coordinates are reported in GRCh37.

**Supplementary Table 18. Variant membership of variance-associated credible sets**. Variants comprising the single-trait credible sets identified for genome-wide significant CMR variance associations. The table reports locus and credible-set identifiers, variant coordinates and alleles and posterior inclusion probabilities. Genomic coordinates are reported in GRCh37.

**Supplementary Table 19. SNP-based heritability estimates for CMR variance associations**. LD Score Regression estimates for variance-association statistics across 22 CMR-derived phenotypes. *h*^2^ denotes SNP-based heritability; SE denotes standard error. The table additionally reports the LD Score Regression intercept and associated standard error and *P* value.

**Supplementary Table 20. GTEx tissue-specific stratified LD Score Regression results for variance CMR associations**. Results are shown for all 53 GTEx tissue annotations across each of the 22 CMR phenotypes. The table reports annotation coefficients, standard errors, *Z* scores and one-sided coefficient *P* values. FDR *q* values were calculated by Benjamini–Hochberg correction across the 53 annotations within each phenotype.

**Supplementary Table 21. HeartMap cardiac cell-type stratified LD Score Regression results for variance CMR associations**. Results are shown for all 14 HeartMap cardiac cell-type annotations across each of the 22 CMR phenotypes. The table reports annotation coefficients, standard errors, *Z* scores and one-sided coefficient *P* values. FDR *q* values were calculated by Benjamini–Hochberg correction across the 14 annotations within each phenotype.

**Supplementary Table 22. Developmental cardiac enhancer stratified LD Score Regression results for variance CMR associations**. Partitioned heritability results are shown for 54 Heart Hub sample-by-state annotations across each of the 22 CMR phenotypes. The table reports annotation composition, heritability enrichment and annotation coefficients. One-sided positive-tail coefficient *P* values were used for Benjamini–Hochberg correction across the 54 annotations within each phenotype.

**Supplementary Table 23. Associations of additive CMR polygenic scores with echocardiographic phenotypes in Penn Medicine Biobank**. Associations of polygenic scores for five CMR phenotypes with corresponding clinically acquired echocardiographic measurements. The table reports analysis sample size, standardized regression coefficient, standard error, 95% confidence interval, *P* value and model R^2^ from linear regression models adjusted for age and sex.

**Supplementary Table 24. Associations of additive and variance CMR polygenic scores with cardiovascular disease in Penn Medicine Biobank**. Associations of additive PGSs and variance PGSs for five CMR traits with dilated cardiomyopathy, any cardiomyopathy, heart failure, atrial fibrillation and ventricular tachycardia. Additive and variance scores were included simultaneously in logistic regression models adjusted for age and sex. Odds ratios, 95% confidence intervals, *P* values and FDR *q* values are reported.

**Supplementary Table 25. Associations of variance CMR polygenic scores with variability in echocardiographic phenotypes**. Variance-PGS validation results for five CMR traits using corresponding extreme echocardiographic measurements in Penn Medicine Biobank. QUAIL models included age, sex and the corresponding additive PGS as covariates. The table reports sample size, effect estimate, standard error, 95% confidence interval, *P* value and FDR *q* value.

**Supplementary Table 26. Replication of individual CMR variance QTLs in Penn Medicine Biobank**. Results for six prespecified genome-wide significant vQTL lead variants with corresponding echocardiographic phenotypes. Analyses were performed in participants of European genetic ancestry using QUAIL. The table reports effect estimates, standard errors, 95% confidence intervals and *P* values, direction concordance with UK Biobank and Bonferroni significance at *P* <0.0083.

**Supplementary Table 27. Additive–variance colocalization at CMR variance-associated regions**. Summary of HyPrColoc analyses jointly evaluating variance associations and additive associations across the 22 CMR traits at the 15 merged variance-associated regions. The table reports overlapping or nearby additive associations, mixed additive–variance clusters, posterior probabilities, candidate variants and traits represented within the strongest mixed cluster.

### Supplementary Figures

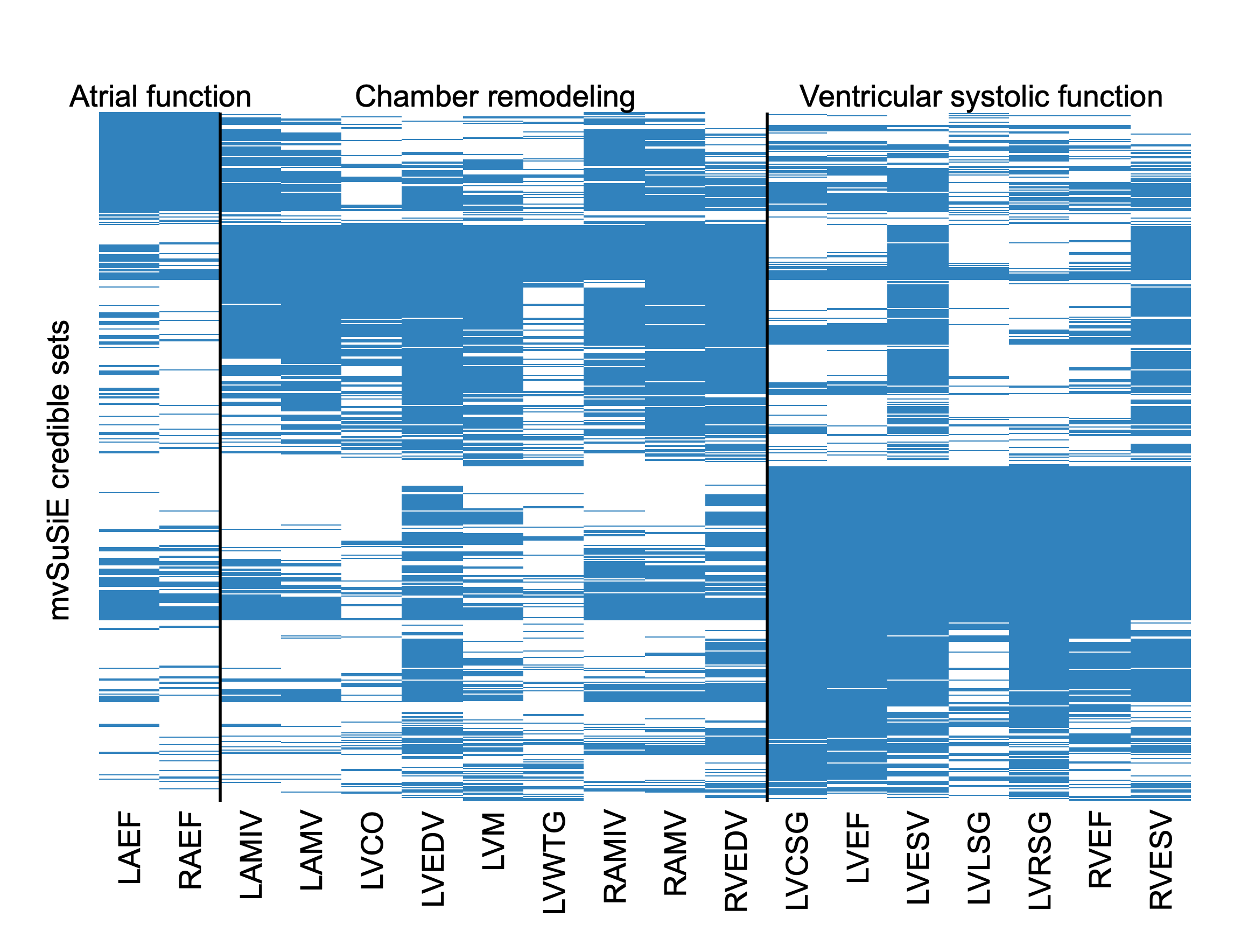

**Supplementary Figure 1. Phenotypic support of multivariate fine-mapped cardiac genetic signals.** Binary matrix showing phenotypic support for the 669 credible sets identified by mvSuSiE across the 18 CMR phenotypes included in multivariate fine-mapping. Rows represent credible sets and columns represent phenotypes; blue cells indicate support for the corresponding phenotype at a single-effect local false sign rate ≤0.01. Phenotypes are ordered according to the three communities identified by Barber bipartite modularity maximization, corresponding broadly to atrial function, chamber remodeling and ventricular systolic function. Credible sets are ordered to display their patterns of phenotypic support within this partition. The observed phenotype partition had Barber bipartite modularity *Q* = 0.181, exceeding that observed in 10,000 randomized networks preserving credible-set and phenotype degree distributions (one-sided empirical *P* = 1.0 × 10^−4^); the same phenotype partition was recovered in all 10,000 random reorderings of the observed network.

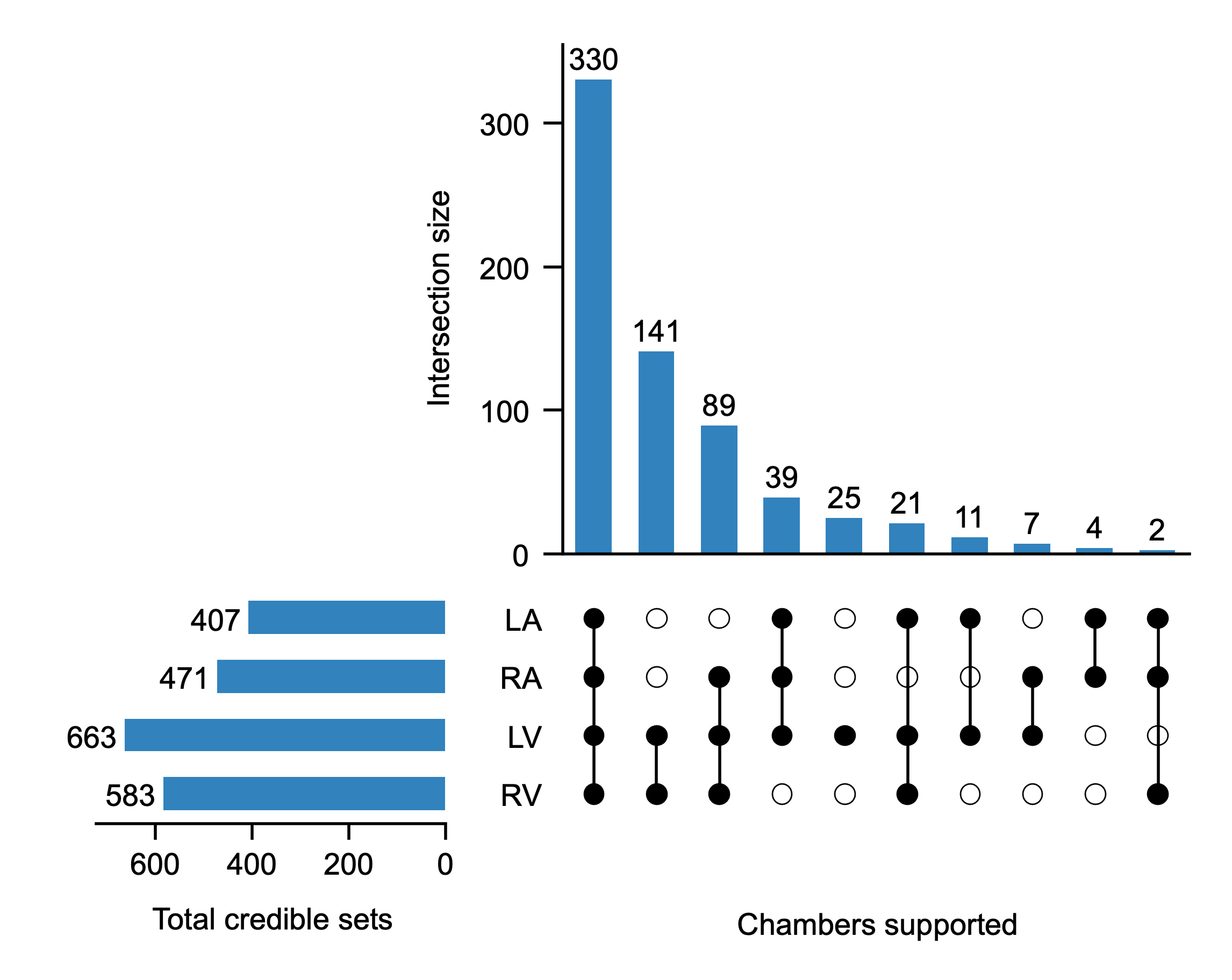

**Supplementary Figure 2. Cardiac-chamber distribution of multivariate fine-mapped genetic signals.** UpSet plot showing the cardiac chambers supported by the 669 mvSuSiE credible sets. Horizontal bars indicate the total number of credible sets supported in each chamber, and vertical bars indicate the number supported by each observed combination of chambers. Chamber support was defined by the presence of at least one phenotype from the corresponding chamber with a single-effect local false sign rate ≤0.01. LA, left atrium; RA, right atrium; LV, left ventricle; RV, right ventricle.

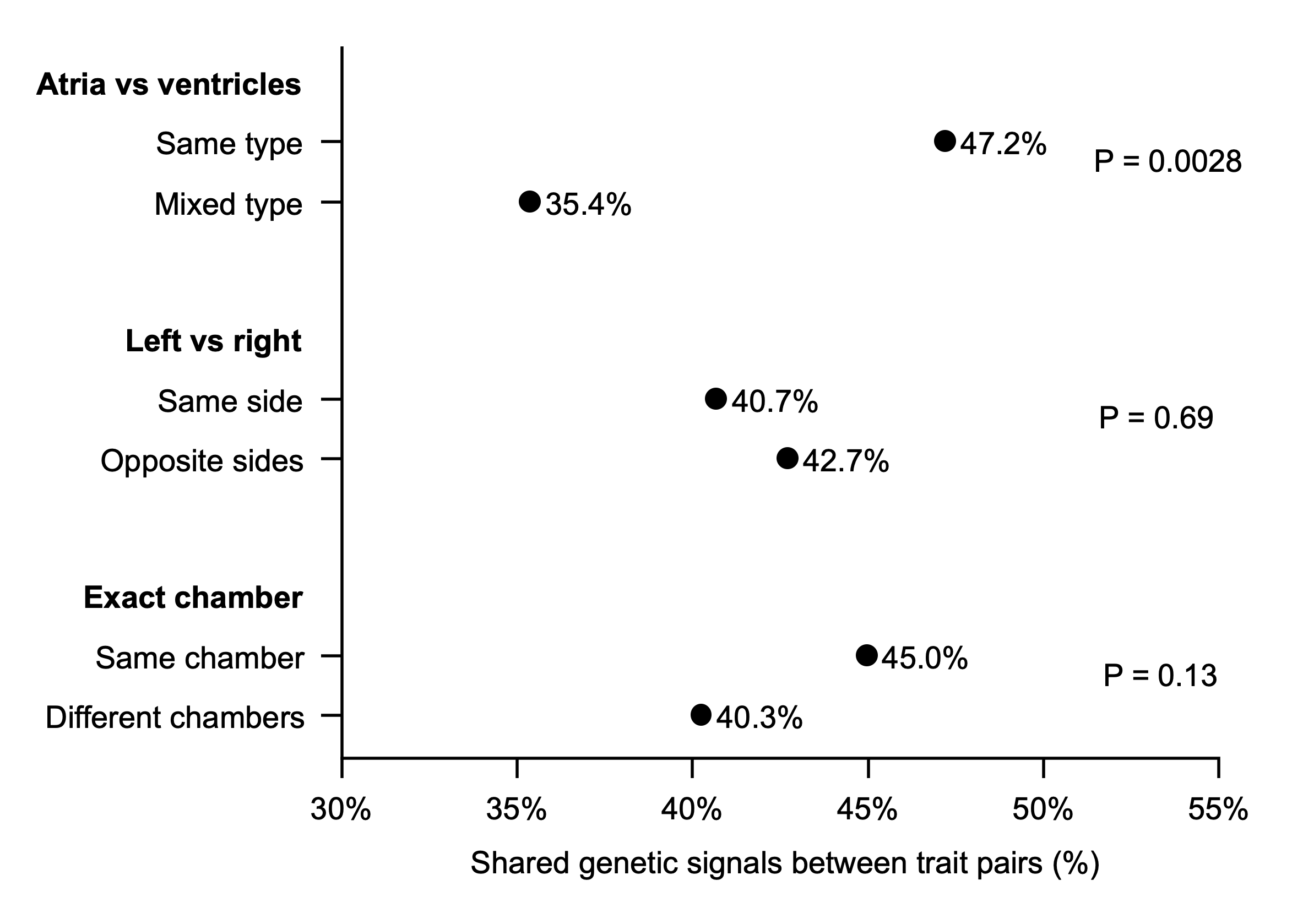

**Supplementary Figure 3. Anatomical organization of cross-trait genetic sharing.** Mean pairwise credible-set sharing according to prespecified anatomical classifications of the 18 CMR phenotypes. Sharing between each pair of phenotypes was quantified using the Jaccard index, defined as the number of credible sets supporting both phenotypes divided by the number supporting either phenotype. Mean sharing was compared between phenotype pairs within the same versus different atrial or ventricular compartments, left–right sides and individual cardiac chambers. Statistical significance was evaluated using 10,000 permutations of anatomical labels across phenotype nodes while preserving the observed credible-set sharing network and the number of phenotypes assigned to each group. One-sided empirical *P* values are shown. Mean sharing was greater within atrial or ventricular compartments than between compartments (47.2% versus 35.4%, *P* = 0.0028), whereas no significant organization was observed by left–right laterality (40.7% versus 42.7%, *P* = 0.69) or individual cardiac chamber (45.0% versus 40.3%, *P* = 0.13).

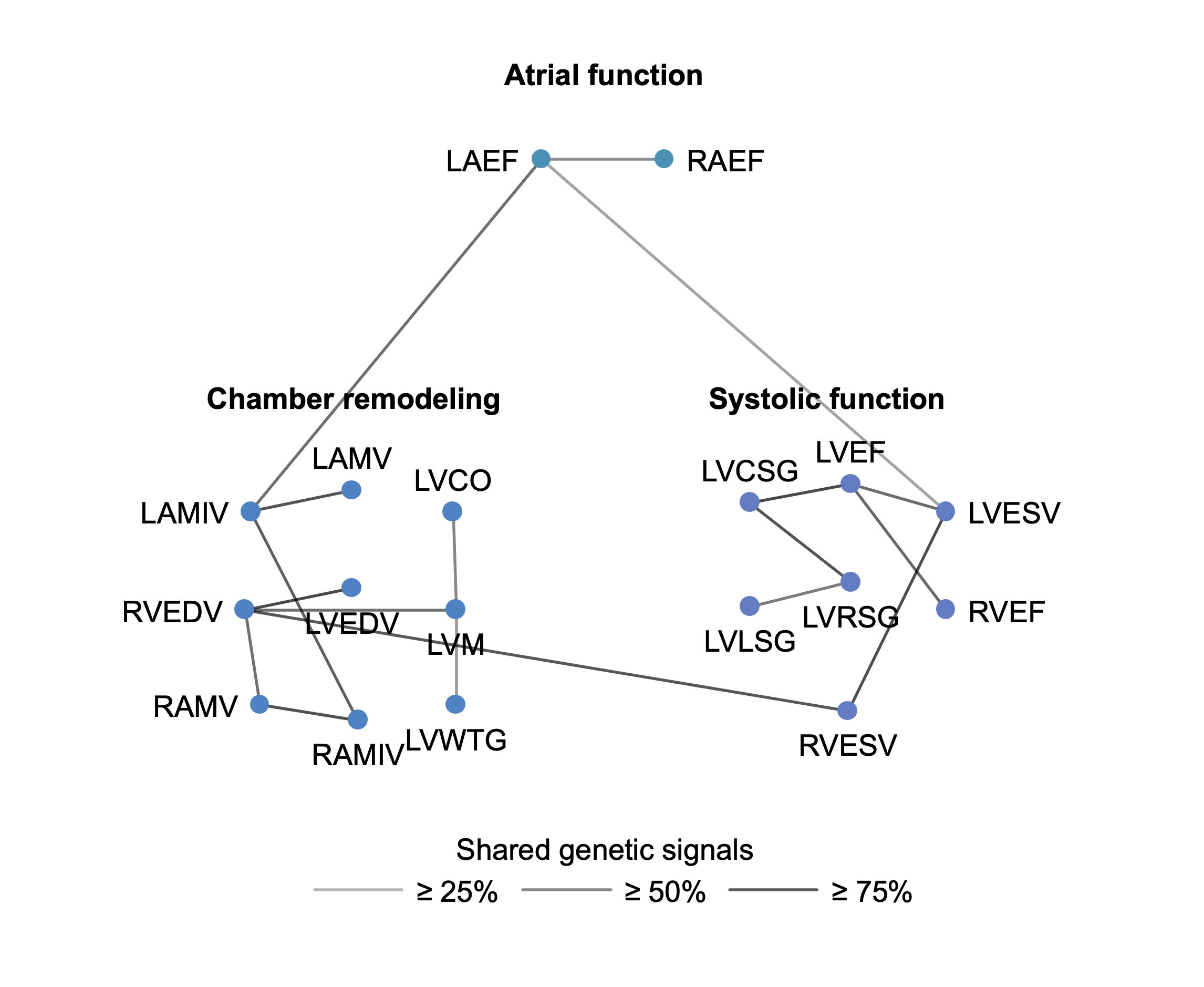

**Supplementary Figure 4. Network organization of genetic sharing across cardiac phenotypes.** Network representation of pairwise credible-set sharing among the 18 CMR phenotypes included in multivariate fine-mapping. Nodes represent phenotypes and are arranged according to the three phenotype communities identified by Barber bipartite modularity maximization, corresponding broadly to atrial function, chamber remodeling and ventricular systolic function. Edges connect phenotype pairs with Jaccard similarity ≥25%, and edge shading denotes increasing levels of credible-set sharing at thresholds of ≥25%, ≥50% and ≥75%.

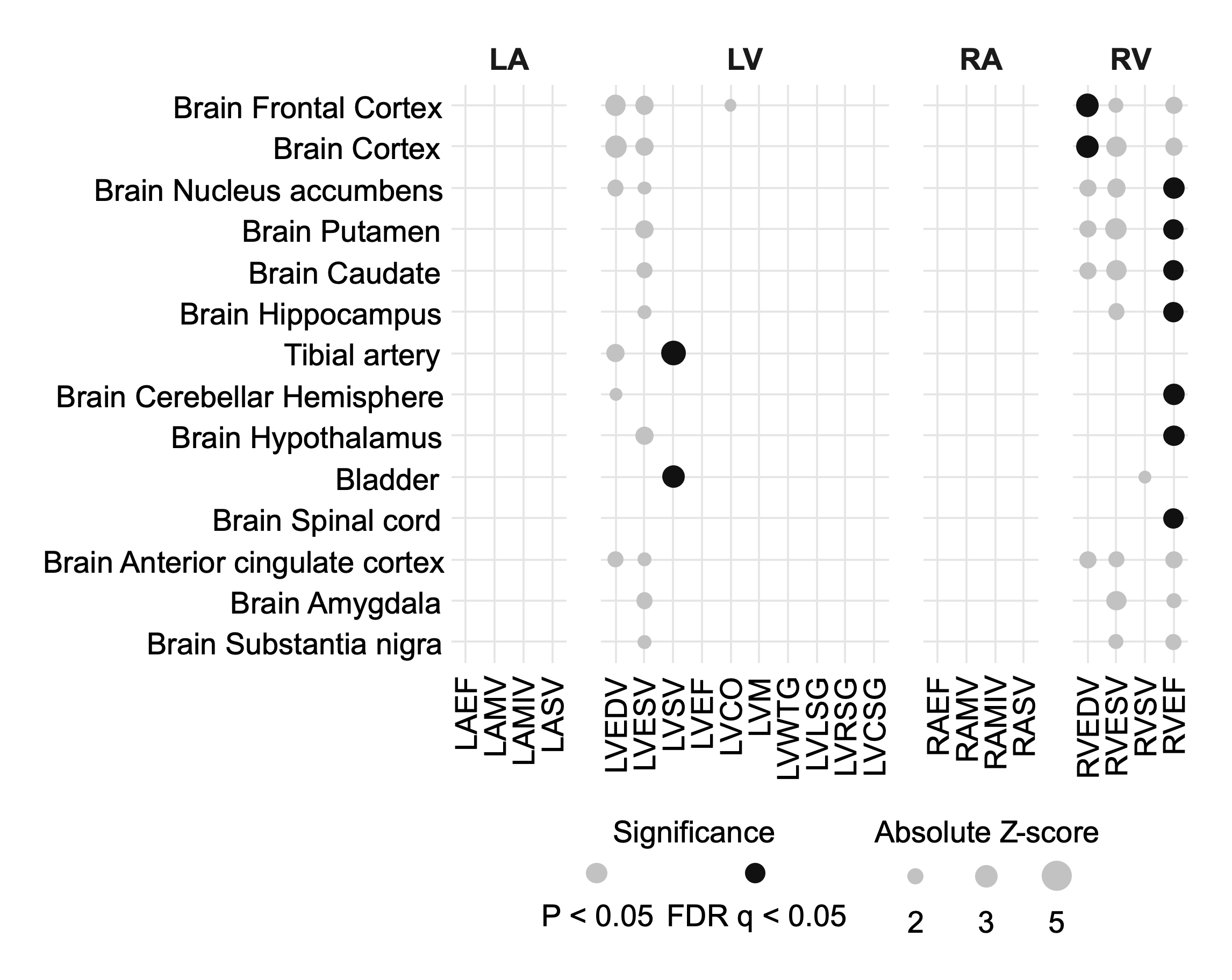

**Supplementary Figure 5. GTEx tissue-specific enrichment of variance-associated CMR genetic effects**. Heatmap of stratified LD Score Regression results for variance-association statistics across 22 CMR phenotypes and 53 GTEx tissue annotations. Cells show annotation coefficient *Z* scores, calculated as coefficient divided by its standard error. FDR significance was determined by Benjamini–Hochberg correction across the 53 annotations within each phenotype; marked cells indicate FDR *q* < 0.05.

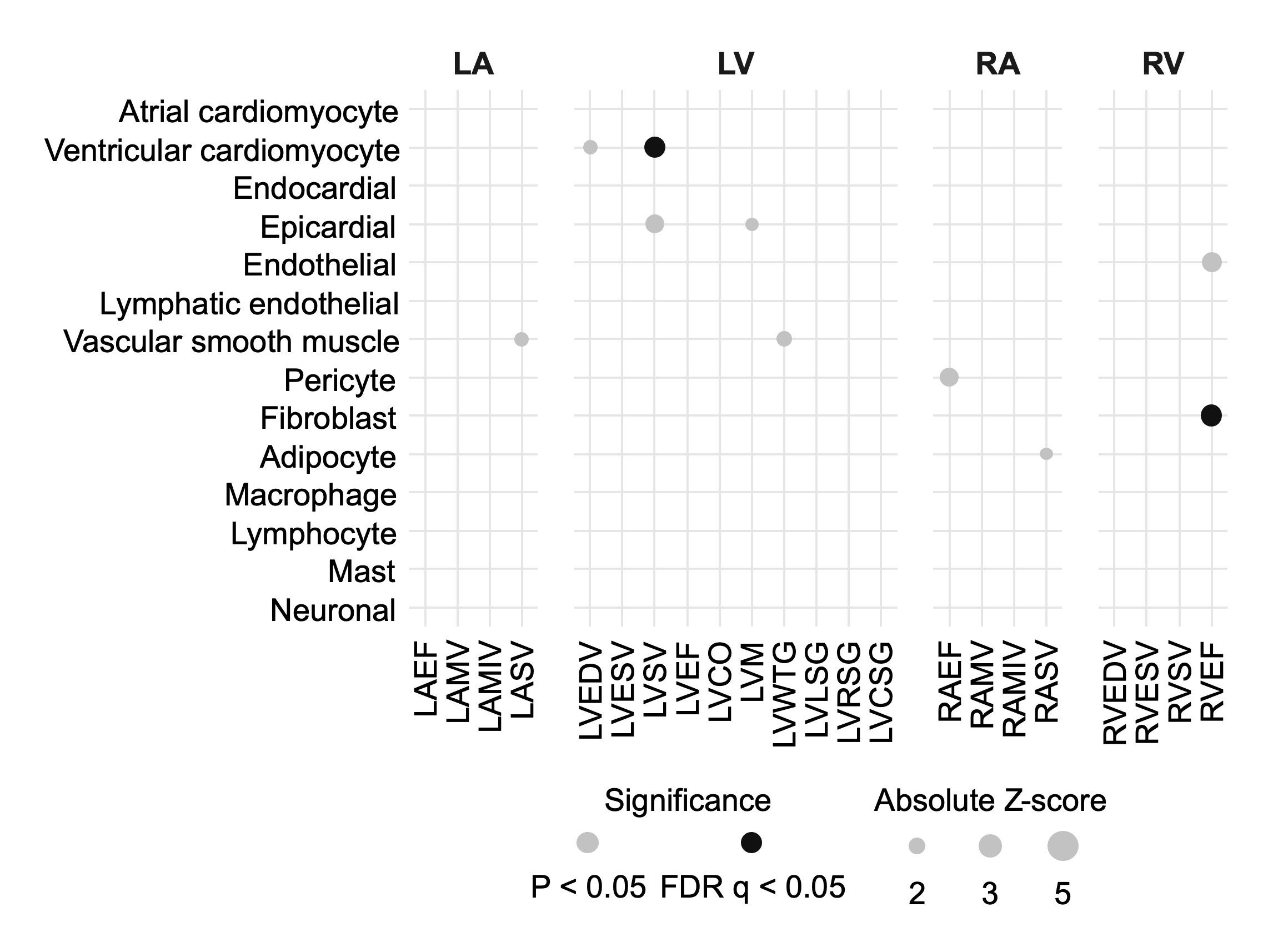

**Supplementary Figure 6. Cardiac cell-type enrichment of variance-associated CMR genetic effects**. Heatmap of stratified LD Score Regression results for variance-association statistics across 22 CMR phenotypes and 14 HeartMap cardiac cell-type annotations. Cells show annotation coefficient *Z* scores, calculated as coefficient divided by its standard error. FDR significance was determined by Benjamini–Hochberg correction across the 14 cell-type annotations within each phenotype; marked cells indicate FDR *q* < 0.05.

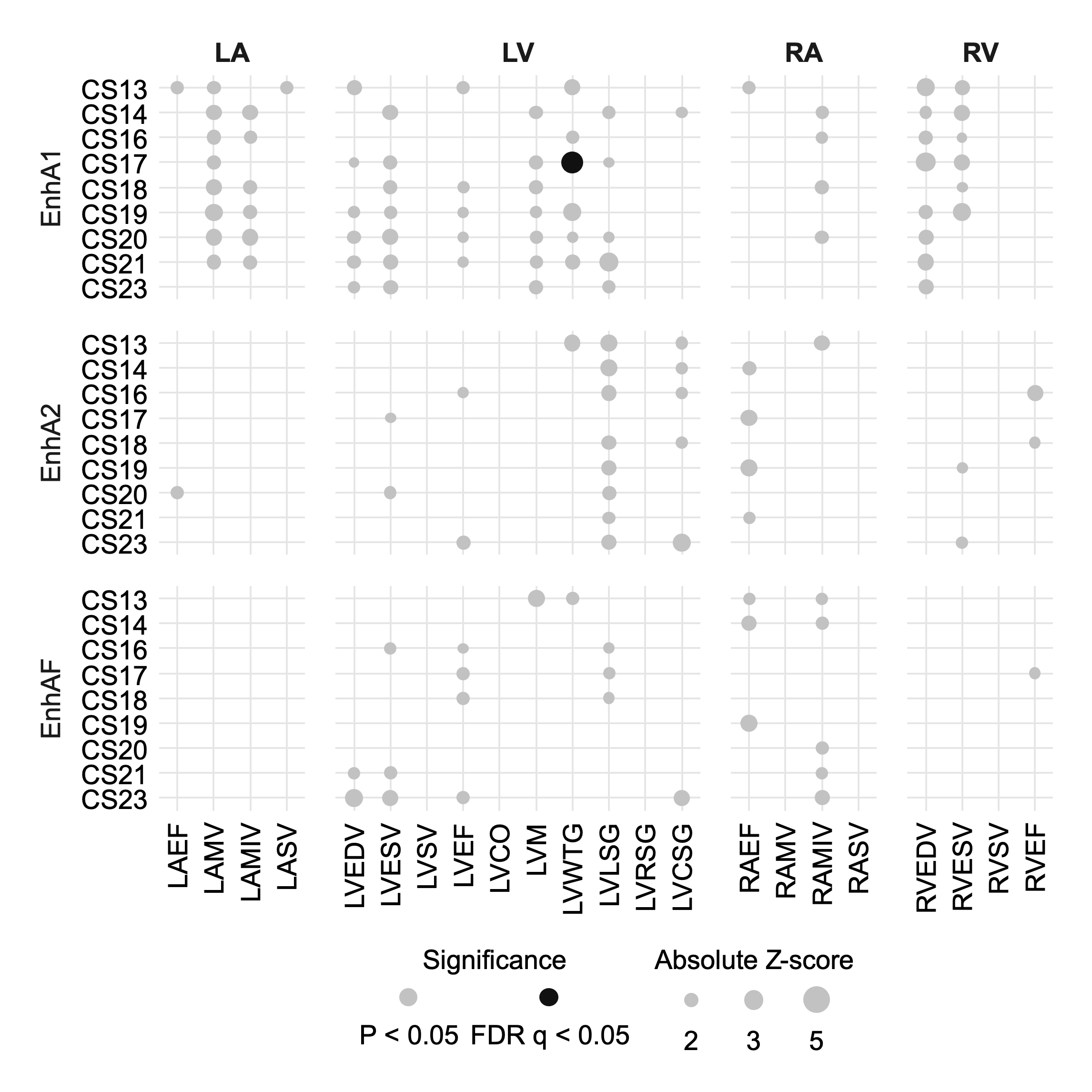

**Supplementary Figure 7. Developmental cardiac enhancer enrichment of variance-associated CMR genetic effects**. Heatmap of stratified LD Score Regression results for variance-association statistics across 22 CMR phenotypes and 54 Heart Hub developmental cardiac enhancer annotations, comprising 18 developmental heart samples and the EnhA1, EnhA2 and EnhAF enhancer states. Cells show annotation coefficient *Z* scores. One-sided positive-tail coefficient *P* values were subjected to Benjamini–Hochberg correction across the 54 annotations within each phenotype; marked cells indicate FDR *q* < 0.05.

### Supplementary Note: Penn Medicine BioBank Banner Author List and Contribution Statements

##### PMBB Leadership Team

Daniel J. Rader, M.D., Marylyn D. Ritchie, Ph.D.

**Contribution:** All authors contributed to securing funding, study design and oversight. All authors reviewed the final version of the manuscript.

##### Patient Recruitment and Regulatory Oversight

JoEllen Weaver, Nawar Naseer, Ph.D., M.P.H., Giorgio Sirugo, M.D., Ph.D., Afiya Poindexter, Yi-An Ko, Ph.D., Kyle P. Nerz

**Contributions:** JW manages patient recruitment and regulatory oversight of study. NN manages participant engagement, assists with regulatory oversight, and researcher access. GS assists with researcher access. AP, YK, KPN perform recruitment and enrollment of study participants.

##### Lab Operations

JoEllen Weaver, Meghan Livingstone, Fred Vadivieso, Stephanie DerOhannessian, Teo Tran, Julia Stephanowski, Salma Santos, Ned Haubein, Ph.D., Joseph Dunn

**Contribution:** JW, ML, FV, SD conduct oversight of lab operations. ML, FV, AK, SD, TT, JS, SS perform sample processing. NH, JD are responsible for sample tracking and the laboratory information management system.

##### Clinical Informatics

Anurag Verma, Ph.D., Colleen Morse Kripke, M.S. DPT, MSA, Marjorie Risman, M.S., Renae Judy, B.S., Colin Wollack, M.S.

**Contribution:** All authors contributed to the development and validation of clinical phenotypes used to identify study subjects and (when applicable) controls.

##### Genome Informatics

Anurag Verma, Ph.D., Shefali S. Verma, Ph.D., Scott Damrauer, M.D., Yuki Bradford, M.S., Scott Dudek, M.S., Theodore Drivas, M.D., Ph.D.

**Contribution:** AV, SSV, and SD are responsible for the analysis, design, and infrastructure needed to quality control genotype and exome data. YB performs the analysis. TD and AV provides variant and gene annotations and their functional interpretation of variants.
